# The potential impact of School Closure Relative to Community-based Non-pharmaceutical Interventions on COVID-19 Cases in Ontario, Canada

**DOI:** 10.1101/2020.11.18.20234351

**Authors:** David Naimark, Sharmistha Mishra, Kali Barrett, Yasin A. Khan, Stephen Mac, Raphael Ximenes, Beate Sander, on behalf of the COVID-19 Modelling Collaborative

**Affiliations:** Institute of Health Policy, Management and Evaluation, University of Toronto, Toronto, Canada; Toronto Health Economics and Technology Assessment (THETA) Collaborative, University Health Network, Toronto, Canada; University Health Network, Toronto, Canada; Sunnybrook Health Sciences Centre, Toronto, Canada; MAP Centre for Urban Health Solutions, Li Ka Shing Knowledge Institute, St. Michael’s Hospital, Toronto, Canada; Department of Medicine, University of Toronto; Institute of Medical Sciences, University of Toronto; Dalla Lana School of Public Health, University of Toronto, Toronto, Canada; Escola de Matemática Aplicada, Fundação Getúlio Vargas, Rio de Janeiro, Brasil; ICES, Toronto, Canada; Public Health Ontario, Toronto, Canada

## Abstract

**Importance:** Resurgent COVID-19 cases have resulted in the re-institution of nonpharmaceutical interventions, including school closure, which can have adverse effects on families. Understanding the impact of schools on the number of incident and cumulative COVID-19 cases is critical for decision-making.

**Objective:** To determine the quantitative effect of schools being open or closed relative to community-based nonpharmaceutical interventions on the number of COVID-19 cases.

**Design:** An agent-based transmission model.

**Setting:** A synthetic population of one million individuals based on the characteristics of the population of Ontario, Canada.

**Participants:** Members of the synthetic population clustered into households, neighborhoods or rural districts, cities or a rural region, day care facilities, classrooms – primary, elementary or high school, colleges or universities and workplaces.

**Exposure:** School reopening on September 15, 2020, versus schools remaining closed under different scenarios for nonpharmaceutical interventions.

**Main Outcome and Measures:** Incident and cumulative COVID-19 cases between September 1, 2020 and October 31, 2020.

**Results:** The percentage of infections among students and teachers acquired within schools was less than 5% across modelled scenarios. Incident case numbers on October 31, 2020, were 4,414 (95% credible interval, CrI: 3,491, 5,382) and 4,740 (95% CrI 3,863, 5,691), for schools remaining closed versus reopening, respectively, with no other community-based nonpharmaceutical intervention; 714 (95%, CrI: 568, 908) and 780 (95% CrI 580, 993) for schools remaining closed versus reopening, respectively, with community-based nonpharmaceutical interventions implemented; 777 (95% credible CrI: 621, 993) and 803 (95% CrI 617, 990) for schools remaining closed versus reopening, respectively, applied to the observed case numbers in Ontario in early October 2020. Contrasting the scenarios with implementation of community-based interventions versus not doing so yielded a mean difference of 39,355 cumulative COVID-19 cases by October 31, 2020, while keeping schools closed versus reopening them yielded a mean difference of 2,040 cases.

**Conclusions and relevance:** Our simulations suggest that the majority of COVID-19 infections in schools were due to acquisition in the community rather than transmission within schools and that the effect of school reopening on COVID-19 case numbers is relatively small compared to the effects of community-based nonpharmaceutical interventions.

**KEY POINTS:** *Question:* With resurgence of COVID-19, reinstitution of school closure remains a possibility. Given the harm that closures can cause to children and families, the expected quantitative effect of school reopening or closure on incident and cumulative COVID-19 case numbers is an important consideration.

*Finding:* Relative to community-based nonpharmaceutical interventions, school closure resulted in a small change in COVID-19 incidence trajectories and cumulative case counts.

*Meaning:* Community-based interventions should take precedence over school closure.

## INTRODUCTION

During the first wave of the COVID-19 pandemic, school closure had been a component of non-pharmaceutical interventions (NPIs) enacted to mitigate the transmission of the SARS-CoV2 virus – largely based on the rationale that it had been effective in delaying or reducing the peak of the 2009 H1N1 influenza epidemic.^1^ It was speculated that school-aged children infected with SARS-CoV2, who are less likely to manifest symptoms,^2^ may unknowingly pass on infections acquired in school to the members of their household and/or individuals at high risk in the community, thereby accelerating the increase in cases and subsequent strain on healthcare resources. In contrast, modelling studies conducted early in the course of the COVID-19 pandemic suggested that school closure was effective when combined with other NPIs, but that the effect of closure *per se* was modest in delaying peak case numbers or reducing the size of the peak.^3–5^

School closures may affect students adversely in terms of loss of access to high-quality instruction, loss of school-based health and social services, and negative effects on physical and emotional well-being.^6–9^ Closures have been shown to result in adverse economic consequences for families due to loss of work hours to care for children and/or increased child-care expenses which predominantly affect lower income households.^6–10^

After closure on March 15, 2020, schools re-opened on September 15, 2020, in Ontario, Canada. As in many jurisdictions, the second wave of COVID-19 in the province has required the reinstitution of community-based NPIs and consideration of school closures. Given the negative consequences of school closure, determining the magnitude of the benefit of this measure in terms of case numbers is critical. The objective of this study is to determine the relative size of the increase in COVID-19 case numbers attributable to school reopening relative to community-based NPIs in Ontario, Canada, using an agent-based modelling approach informed by observed Ontario data.

## METHODS

### Development of a synthetic population representative of Ontario, Canada

We developed a synthetic population of one million individuals representative of the inhabitants of Ontario, Canada (population ∼ 14.5 million), based on data within the Social Policy Simulation Database and Model (SPSD/M, **eMethods**)^11^ developed by Statistics Canada. SPSD/M contains data on Ontario households categorized by rural or urban location, numbers of household members, age and gender of household members, labour force participation and industry. For the synthetic population, households were randomly sampled from the available household types to yield a total of one million hypothetical individuals. Households were then randomly assigned to cities or a rural region according to the SPSD/M urban/rural designation of the associated household type. Within urban settings, households were further randomly assigned to neighborhoods while rural household were assigned to districts.

All members of the synthetic population spent time each day in their households, neighborhoods or districts, and their city or rural region. Additionally, children between 2 and 3 years old were assigned to daycare settings, children between 4 and 13 were assigned to primary/elementary schools, and children between 14 and 17 were assigned to high schools. Each daycare or classroom was assigned a teacher who was randomly sampled from adults in the region whose SPSD/M industry designation was educational. Adults, 18 to 34 years old, could join the workforce or potentially go to college or university (a post-secondary institution). Adults 18 to 34 years old, not enrolled in a post-secondary institution, and those aged 35 to 64 years old could be members of the workforce. Workplaces were characterized by industry type, region, and workplace size. Working age adults were then randomly assigned to workplaces in their region according to their SPSD/M industry designator. Adults, 65 and older, were assumed to be retired from the workforce.

### Agent-based model

The Agent-Based Model for COVID-19 Transmission (ABMCT) was structured to represent the COVID-19 pandemic in Ontario and made use of the synthetic population described above (**eMethods and eFigure 1**). For the COVID-19 first wave, the model was seeded with 150 infectious individuals who were selected from the synthetic population at random. All other individuals were initially susceptible to COVID-19 and could contact and be exposed to infectious individuals within their household, at school, at college or university, at workplaces, in neighborhoods and cities, or rural districts. Exposed individuals may not have been infected and remain susceptible or they may have become infected. Infected individuals were unable to transmit the virus during a four-day latent period, after which, infected individuals became infectious and able to transmit the virus. Infectious individuals may or may not have developed symptoms. The former entered a one-day pre-symptomatic stage followed by a symptomatic stage. The duration of infectiousness for both individuals with and without symptoms was modelled to be 15 days. The proportion of symptomatic cases by ten-year age group was obtained from Ontario’s Case and Contact Management Plus (CCM Plus) database (**eMethods and eTable1**).^12^

**Figure 1.**
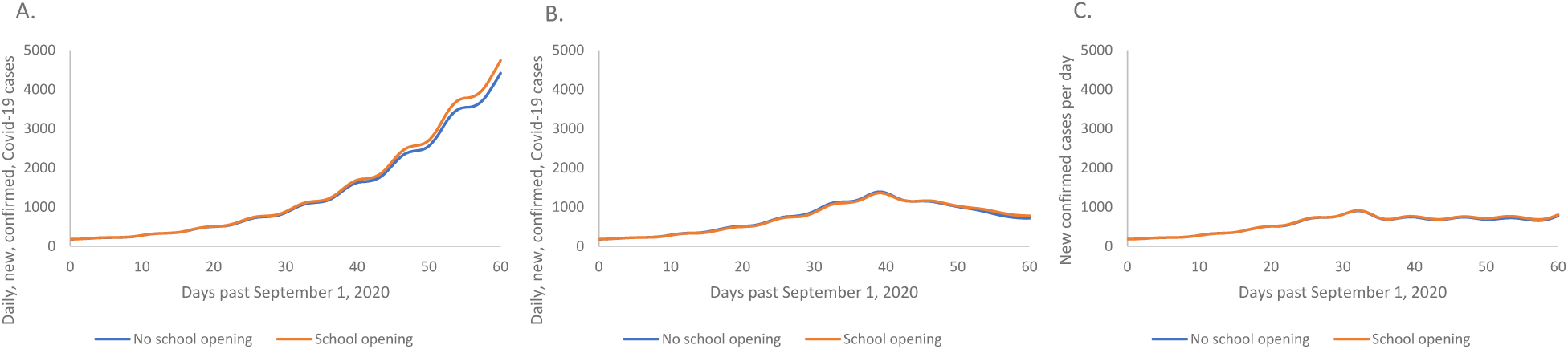
Mean of 100 model replicates for the case that schools had re-opened (orange) or had remained closed (blue) on September 15, 2020. Note, modelled counts subjected to Gaussian kernel smoothing with a bandwidth of 2 days (**eMethods**). **Panel – A**. Comparison of model simulation results for public health measure scenario 1: incident case numbers from September 1 to October 31, 2020, given no public health interventions restricting contacts and/or reducing the probability of transmission of COVID-19 between contacts had been implemented (that is, if the trends in the rise of new cases in September had been allowed to persist through October). **Panel – B**. Comparison of model simulation results for public health measure scenario 2: incident case numbers from September 1 to October 31, 2020, given implementation of public health interventions restricting contacts and/or reducing the probability of transmission of COVID-19 between contacts had been implemented (that is, if the trends in the rise of new cases in September *had not been allowed* to persist through October). **Panel – C**. Comparison of model simulation results for public health measure scenario 3: incident case numbers from September 1 to October 31, 2020, given that the slowing of the rate of growth of cases observed from 1 – 15, October, 2020, to 0.8% per day persisted until October 31, 2020.

Symptomatic individuals could seek health care, be confirmed as cases, and either be admitted to hospital or sent home to quarantine until recovered. Symptomatic individuals who did not seek health care could self-isolate until recovery. Self-isolated or quarantined individuals could still transmit SARS-CoV2 to their household members. A proportion of asymptomatic cases would be detected through contact tracing, reported as confirmed cases, and quarantined until recovery. Symptomatic individuals who neither seek health care nor testing, and undetected asymptomatic individuals would be free to transmit virus in household and non-household settings. The probability of detection, i.e. of an infectious individual being a confirmed case, during the first wave of COVID-19 in Ontario was estimated, separately for symptomatic and asymptomatic cases, via the cumulative number of confirmed cases until June 9, 2020, and the seroprevalence of antibodies for SARS-CoV2 reported by Public Health Ontario until that date.^13^ Death outside of hospital, such as in a long-term care (LTC) facility, was not considered in the current iteration of the model nor was nosocomial transmission within hospitals.

Susceptible individuals were modelled to be potentially infected via close contact with an infectious individual. Mixing of individuals was assumed to occur randomly within the settings mentioned previously based on the average number of close contacts per day prior to the institution of NPIs in March and April 2020 from a contact survey, the CONNECT study^14^ **(Table 1, eMethods, eFigures 2 - 4)**. We calibrated the model to observed COVID-19 first-wave data **(Table 1, eMethods, eTable 2, and eFigures 5-7)** and second-wave data **(Table 2, eMethods and eFigures 8 – 13)**.

**Table 1.**
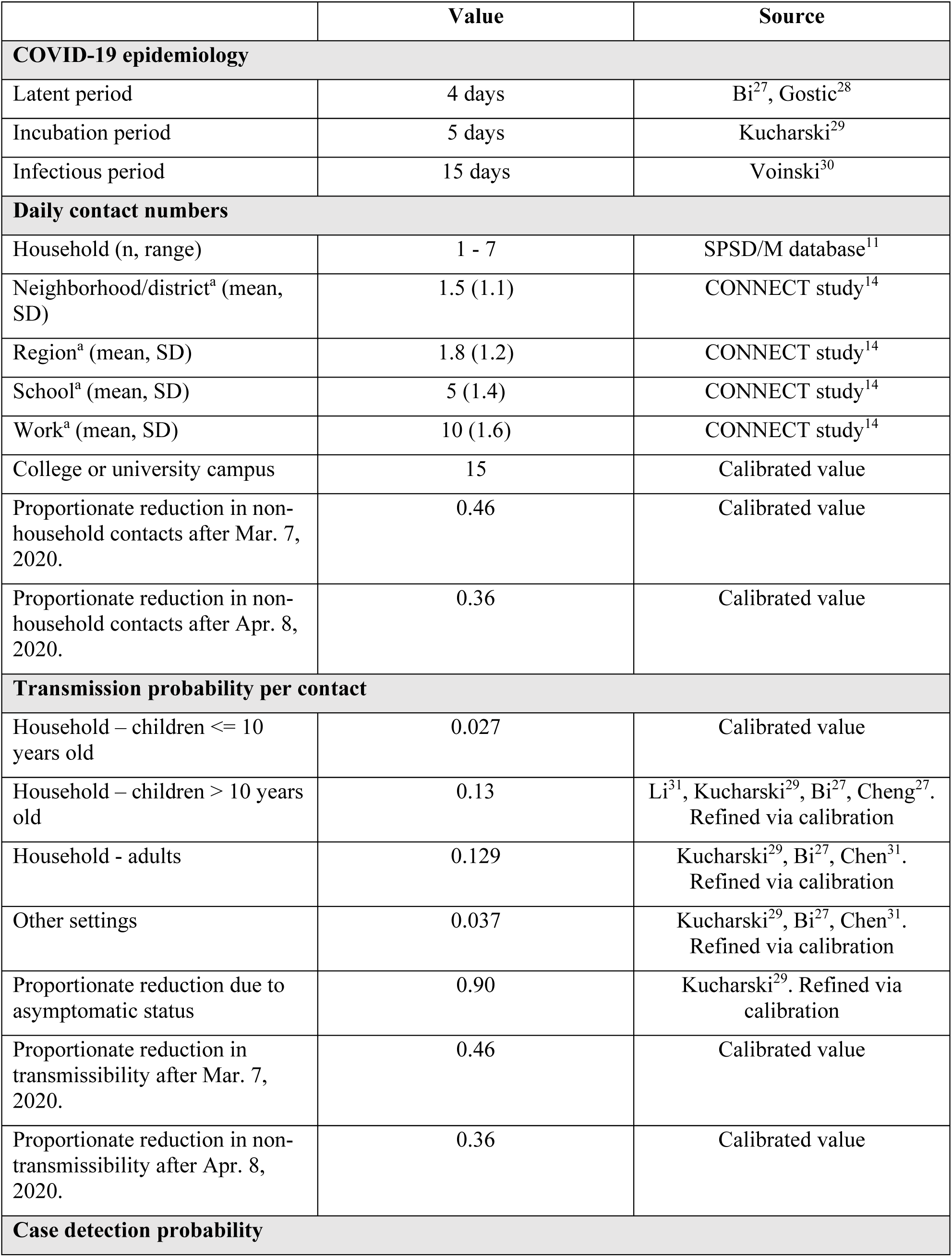
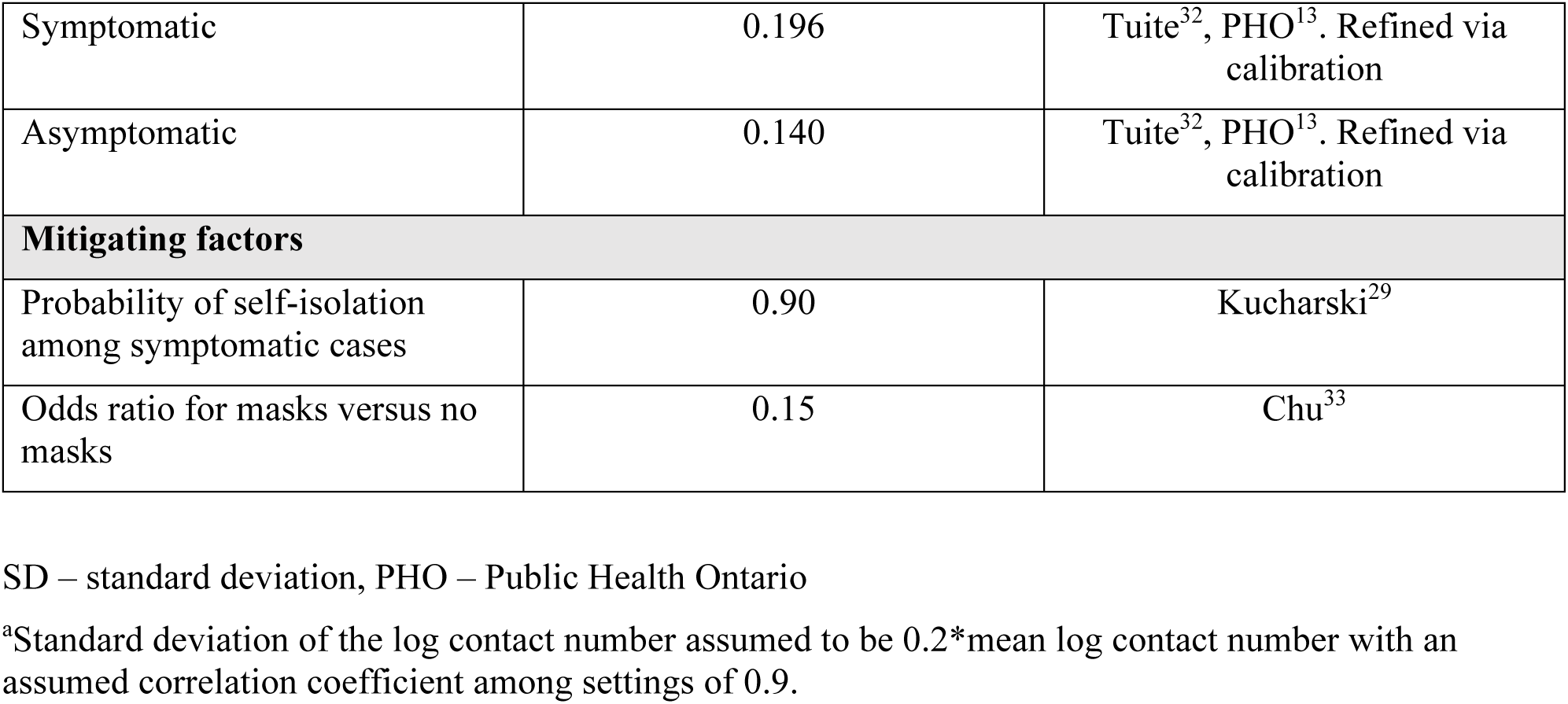
Key parameters for ABMCT - first wave.

**Table 2.** Key parameters for ABMCT 1.0 - second wave.

|  | Value | Source |
| --- | --- | --- |
| **Daily contact numbers** |  |  |
| Proportionate reduction in non-household contacts until Oct 1., 2020, compared to pre-pandemic. | 0.81 | Wang<sup>34</sup> |
| Proportionate reduction in non-household contacts after Oct 1., 2020, compared to pre-pandemic.<sup>a</sup> | 0.4 | Re-calibrated value |
| **Transmission probability per contact** |  |  |
| Household – children <= 10 years old | 0.024 | Re-calibrated value |
| Household – children > 10 years old | 0.116 | Li<sup>31</sup>, Kucharski<sup>29</sup>, Bi<sup>27</sup>, Cheng<sup>35</sup>. Refined via recalibration |
| Household - adults | 0.115 | Kucharski<sup>29</sup>, Bi<sup>27</sup>, Cheng<sup>35</sup>. Refined via re-calibration |
| Other settings | 0.033 | Kucharski<sup>29</sup>, Bi<sup>27</sup>, Cheng<sup>35</sup>. Refined via re-calibration |
| Proportionate reduction in non-transmissibility after Oct 1., 2020, compared to pre-pandemic.* | 0.5 | Re-calibrated value |
| **Case detection probability** |  |  |
| Symptomatic | 0.287 | Tuite<sup>32</sup>, PHO<sup>13</sup>. Refined via re-calibration |
| Asymptomatic | 0.187 | Tuite<sup>32</sup>, PHO<sup>13</sup>. Refined via re-calibration |

PHO – Public Health Ontario. Note that only parameters different from those in Table 1 are listed.
<sup>a</sup>In scenarios in which public health restrictions were imposed on October 1, 2020.

**Figure 2.**
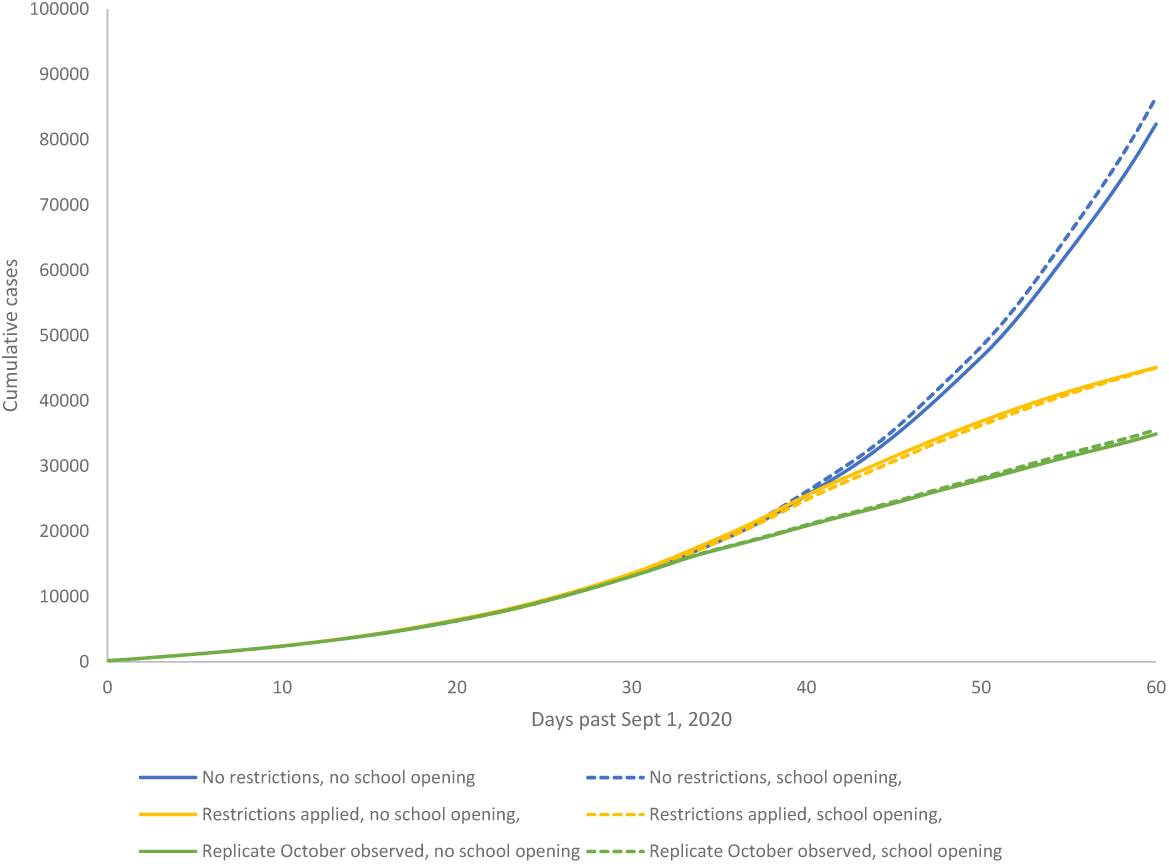
Comparison of model simulations for the cumulative number of COVID-19 cases in Ontario between September 1 and October 31, 2020, among the six modelled scenarios. Mean of 100 model replicates for the case that no restrictions were implemented and schools had not opened (blue); no restrictions were implemented and schools had re-opened (dotted blue); restrictions were implemented and schools had not opened (yellow); restrictions were implemented and schools had opened (dotted yellow); the slowing of the growth of new case numbers observed from October 1 – 15, 2020, had persisted until October 31, 2020, had schools not re-opened (green) or had re-opened (dotted green). Modelled counts subjected to Gaussian kernel smoothing with a bandwidth of 2 days (**eMethods**).

### Daycare and school opening scenarios

We simulated two school opening scenarios (A and B, **eTable 3**). First, we modelled a counterfactual scenario in which schools did not reopen on September 15, 2020 (scenario A). Second, we modelled a scenario in which schools re-opened on September 15, 2020, (scenario B) but with several measures in place to limit within-school transmission of COVID-19: i) daycare centres were capped at 10 children, primary and elementary class sizes were capped at 23, and high school classes were capped at 15 students; ii) students remained in their assigned classrooms for the school day rather than moving among classrooms; iii) universal masking was in place; iv) in designated high schools, in urban areas, students attended school only on alternate weekdays; and v) if more than two confirmed cases of COVID-19 occurred in a daycare or classroom less than two weeks apart, the daycare or classroom was closed for 14 days with the children in the class excluded from school rather than moved to another classroom (**eTable 3**).

### Non-pharmaceutical intervention scenarios

We modelled three community-based NPI scenarios (1,2 and 3, **eTable 3**) in Ontario at the beginning of October 2020, in response to rising confirmed daily case incidence from 185 on September 1^st^ to 675 on September 30^th^ (**eFigures 8 – 10**). First, we modelled a scenario in which no additional NPIs were enacted so that the increase in case numbers observed in September continued, unabated, until October 31, 2020. Second, we modelled a scenario in which community-based NPIs were enacted on October 1, 2020, that resulted in a reduction in contacts outside of households to 40% of the value observed pre-pandemic, including closing of workplaces to non-essential workers, and a reduction in the probability of transmission per contact of 50% compared to the re-calibrated values within household and other settings. Third, we modelled a scenario in which the growth of new infections was limited to 0.8% per day replicating the reduced growth of confirmed cases in the CCM database for the first 15 days of October (**eTable3**).

The two school reopening and three community-based NPI scenarios created 6 combinations, 1A to 3B (**eTable 3**). One hundred repetitions of the ABMCT were run for each combination. All analyses were conducted with TreeAge Pro 2020 R2 (Williamstown, Mass.).

## RESULTS

### All results are presented on a provincial scale

The median (inter-quartile range, IQR) ***number of classroom closures*** for scenarios involving school reopening, i.e., lack of community-based NPI implementation (scenario 1B), NPI implementation (scenario 2B), and replication of observed COVID-19 case counts in early October (scenario 3B), were 1,726 (421), 1,051 (337) and 587 (178), respectively (**eTable 4, eFigures 14 and 15**), with the majority of closures occurring in primary and elementary schools. For teachers and students, median (IQR) numbers of infections transmitted within schools were 493 (134), 421 (109) and 167 (58) while the numbers acquired in the community were 15,892 (1,842), 9,643 (1,407) and 7,265 (848) corresponding to percentages of total infections acquired in schools of 3.15% (0.76%), 4.19% (0.80%) and 2.37% (0.84%), respectively (**eTable 4, eFigures 14 and 15**).

When community-based NPIs were not enacted, the ***number of daily, new, confirmed cases on October 31, 2020***, were 4,414 (95% credible interval, CrI: 3,491; 5,382) with schools closed (scenario 1A), versus 4,740 (95% CrI 3,863; 5,691) with schools reopened (scenario 1B), with an increase of cases of due to school reopening of 326 (95% CrI:196; 456) (**Figure 1 - panel A, eFigures 16 and 17**). If community-based NPIs were implemented, the number of daily, new, confirmed cases on October 31, 2020, were 714 (95% CrI: 568; 908) with schools closed (scenario 2A), versus 780 (95% CrI 580; 993) with schools reopened (scenario 2B), with an increase of cases of due to school reopening of 66 (95% CrI: 40; 92) (**Figure 1 – panel B, eFigures 18 and 19**). For the two scenarios where the change in the growth of observed case counts in early October, 2020, was replicated, the number of daily, new, confirmed cases on October 31, 2020, were 777 (95% CrI: 621; 993) with schools closed (scenario 3A), versus 803 (95% CrI 617; 990) with schools reopened (scenario 3B), with an increase of cases of due to school reopening of 26 (95% CrI: 0; 52) (**Figure 1 – panel C, eFigures 20 and 21**).

The average, ***cumulative number of confirmed COVID-19 cases, by October 31, 2020***, were 82,372 and 86,507 with schools closed (scenario 1A) and reopened (scenario 1B), respectively, with no community-based NPIs enacted, a difference of 4,135 cases (**Figure 2**). The cumulative cases were 45,112 and 45,068 with schools closed (scenario 2A) and reopened (scenario 2B), respectively, when community-based NPIs were implemented, a difference of 54 cases (**Figure 2**). The cumulative cases were 34,911 and 35,581 with schools closed (scenario 3A) and reopened (scenario 3B), when the decrease in the growth of observed case counts in early October, 2020, was replicated, a difference of 670 cases (**Figure 2**). The mean difference in cumulative COVID-19 cases by October 31, 2020, for the scenarios in which community-based NPIs were not implemented (1A and 1B) versus scenarios in which they were (2A and 2B) was 39,355 cases (**eFigure 22**). In contrast, the mean difference in cumulative COVID-19 cases by October 31, 2020, for the scenarios in which schools were reopened (1B and 2B) versus scenarios in which they were not (1A and 2A) was 2,040 cases (**eFigure 22**).

## DISCUSSION

Our simulations indicate that school reopening compared to the counterfactual situation where they remained closed on September 15, 2020, leads to a modest increase in incident and cumulative COVID-19 cases across three community-based NPI implementation scenarios in Ontario, Canada, between September 1 and October 31, 2020. The magnitude of the increase related to school reopening was driven mostly by rates of community spread with children and teachers bringing SARS-CoV2 infections into the school rather than transmitting them within schools. Thus, community-based NPIs directed at reducing contacts, such as restricting gatherings, limiting workplaces to essential workers, and reducing transmission between contacts, for example by requiring mask wearing, had a much larger effect in our simulations on reducing incident or cumulative COVID-19 cases than keeping schools closed versus reopening them. It is important to note that our model assumed that measures would be taken in schools to limit spread of the virus by reducing high school class sizes, having students remain in the same classroom rather than moving among them, requiring universal mask wearing, and closing classrooms when more than two confirmed SARS-CoV2 infections occurred within any two week period.

The results of our simulations are in agreement with a meta-analysis of COVID-19 pandemic data in Hong Kong, Singapore and mainland China conducted by Viner at al.^15^ who found that school closures did not materially contribute to the prevention of population spread. Likewise, in Denmark, the first country in Europe to reopen schools, the incidence of COVID-19 continued to decline after reopening of day care facilities and elementary schools in mid-April and middle and high schools in May, 2020, albeit with stringent physical distancing measures in place.^16^ Our results were also similar to several prior modelling studies.^3–5^ However, these were conducted early in the course of the pandemic when precise information regarding transmission probability between cases and/or when observed data required for model calibration was unavailable and/or did not distinguish between infections acquired in school versus those acquired in the community and imported into schools. Our results bolster the prior studies but considered events later in the course of the pandemic, at the traditional time of school reopening in Canada, and made use of accumulated, observed, calibration data.

Our results, showing that the majority of infections in schools are due to community acquisition rather than transmission within schools, are congruent with numerous contact tracing studies of secondary infections in schools conducted in many jurisdictions which show very infrequent onward transmission within schools.^17–23^ For example, in a systematic review, Public Health Ontario^24^ synthesized four studies of outbreak investigations which showed that 28 index cases among students and teachers lead to 2,092 close contacts from which there were only two transmissions. Forbes et al.^25^ in an analysis of health services data in the United Kingdom, involving more than nine million adults less than 65 years old, showed that living with a child 11 years old or younger was not associated with increased SARS-CoV2 infections, or COVID-19 ward or ICU admissions. Furthermore, data obtained in the United Kingdom showed a strong correlation between the frequency of school outbreaks and regional COVID-19 incidence.^26^

The relatively modest reduction in COVID-19 cases derived from keeping schools closed should be balanced against the adverse effects on children and families. School closure may interfere with educational advancement, prevent access to school-based health and social programs, reduce physical activity among children, and negatively affect household finances particularly among low income households.^10^

Our analyses have limitations. Like all models, our agent-based model is a simplification of complex, dynamic patterns of interactions and viral transmission in real populations. For example, although close contacts may occur among students, particularly for adolescents, just before and after school hours, this was not explicitly modelled. However, this mixing was partially captured in the close contacts among individuals in neighborhoods or rural districts. Modelled school opening and NPI scenarios may not precisely reflect the actual implementation of public health policy in Ontario but have the advantage of quantitatively determining the effect of school reopening across a range of community-based NPI possibilities with robust qualitative results. Also, by not being overly jurisdiction-specific, the results of the model may be broadly applicable to regions similar to Ontario, in terms of school systems, populations, and the economy. Latent, incubation, and infectious periods were assumed to be fixed rather than drawn from distributions which may have caused an underestimation in the uncertainty of pandemic trajectories but would not have affected the average results. The model did not explicitly consider COVID-19 testing volumes as a potential driver of confirmed case numbers but rather assumed that, over time, testing rates, and therefore case detection probability, would stabilize.

Our analysis has several strengths. The use of a synthetic population allows for realistic linkages of individuals into discrete clusters such as households, classrooms and workplaces unlike other approaches that treat mixing of individuals among age strata in a probabilistic fashion. The agent-based modelling approach allows for so-called super-spreading events, where a small number of individuals have a disproportionately large numbers of close contacts, and for representation of relatively complicated measures such as classroom closure after a certain number of confirmed cases over a particular interval, which would be difficult with other transmission model designs such as compartmental models. Unlike previous transmission models, we had the advantage of being able to calibrate our model to observational data regarding case numbers in Ontario from March through September, 2020. Agent-based modelling allows the source of infections to be determined allowing the disentanglement of infections transmitted in schools versus those acquired in the community and brought into the school setting.

In conclusion, the magnitude of the effect of schools being open on COVID-19 cases is substantially lower than the effect of community-based NPIs. The results of this study suggest that school closure be considered the last resort in the face of a resurgence of COVID-19, and that efforts instead focus on widespread reduction of community transmission.

## Data Availability

Data employed in this analysis is in the public domain

## Acknowledgements

The authors would like to thank Richard Shillington, PhD for help with queries of the Social Policy Simulation Database and Model (SPSD/M) and Al Crosny, PhD for technical help with the transmission model’s software platform.

## REFERENCES

1. Isfeld-Kiely, Harpa; Moghadas S. Effectiveness of School Closure for the Control of Influenza A Review of Recent Evidence.; 2014.

2. Qiu H, Wu J, Hong L, Luo Y, Song Q, Chen D. Clinical and epidemiological features of 36 children with coronavirus disease 2019 (COVID-19) in Zhejiang, China: an observational cohort study. Lancet Infect Dis. 2020;20(6):689–696. doi:10.1016/S1473-3099(20)30198-5

3. Davies NG, Kucharski AJ, Eggo RM, et al. Effects of non-pharmaceutical interventions on COVID-19 cases, deaths, and demand for hospital services in the UK: a modelling study. Lancet Public Heal. 2020;5(7):e375–e385. doi:10.1016/S2468-2667(20)30133-X

4. Koo JR, Cook AR, Park M, et al. Interventions to mitigate early spread of SARS-CoV-2 in Singapore: a modelling study. Lancet Infect Dis. 2020;20(6):678–688. doi:10.1016/S1473-3099(20)30162-6

5. Banholzer N, van Weenen E, Kratzwald B, et al. Impact of non-pharmaceutical interventions on documented cases of COVID-19. medRxiv. January 2020:2020.04.16.20062141. doi:10.1101/2020.04.16.20062141

6. Effler P V., Carcione D, Giele C, Dowse GK, Goggin L, Mak DB. Household responses to pandemic (H1N1) 2009-related school closures, Perth, Western Australia. Emerg Infect Dis. 2010;16(2):205–211. doi:10.3201/eid1602.091372

7. Basurto-Dávila R, Garza R, Meltzer MI, et al. Household economic impact and attitudes toward school closures in two cities in Argentina during the 2009 influenza A (H1N1) pandemic. Influenza Other Respi Viruses. 2013;7(6):1308–1315. doi:10.1111/irv.12054

8. Gift TL, Palekar RS, Sodha S V., et al. Household effects of school closure during pandemic (H1N1) 2009, Pennsylvania, USA. Emerg Infect Dis. 2010;16(8):1315–1317. doi:10.3201/eid1608.091827

9. Epson EE, Zheteyeva YA, Rainey JJ, et al. Evaluation of an Unplanned School Closure in a Colorado School District: Implications for Pandemic Influenza Preparedness. Disaster Med Public Health Prep. 2015;9(1):4–8. doi:10.1017/dmp.2015.3

10. Ontario Agency for Health Protection and Promotion (Public Health Ontario). Negative Impacts of Community-Based Public Health Measures during a Pandemic (e.g., COVID-19) on Children and Families. Toronto; 2020.

11. Statistics Canada. Social Policy Simulation Database and Model (SPSD/M) Version 28.0.1. 2020. https://search2.odesi.ca/#/details?uri=%2Fodesi%2Fspsd-89F0002-E.xml.

12. Ontario Ministry of Health. Integrated Public Health Information Systems (IPHIS).; 2020.

13. Ontario Public Health. COVID-19 Seroprevalence in Ontario: March 27, 2020 to June 30, 2020.; 2020.

14. Brisson, Marc; Gingras, Guillaume; Drolet, Melanie; Laprise J-F. Modélisation de l’évolution de la COVID-19 au Québec. http://www.marc-brisson.net/covid19-response/Epidemiologie-et-modelisation-evolution-COVID-19-au-Quebec_16-octobre-2020.pdf.

15. Viner RM, Russell SJ, Croker H, et al. School closure and management practices during coronavirus outbreaks including COVID-19: a rapid systematic review. Lancet Child Adolesc Heal. 2020;4(5):397–404. doi:10.1016/S2352-4642(20)30095-X

16. Levinson, Meira; Muge, Cevik; Lipsich M. Reopening Primary Schools during the Pandemic. N Engl J Med. 2020;383(10):981–985.

17. Danis K, Epaulard O, Bénet T, et al. Cluster of coronavirus disease 2019 (COVID-19) in the French Alps, February 2020. Clin Infect Dis. 2020;71(15):825–832. doi:10.1093/cid/ciaa424

18. Yung CF, Kam K, Nadua KD, et al. Novel Coronavirus 2019 Transmission Risk in Educational Settings. Clin Infect Dis. 2020;(Xx Xxxx):3–6. doi:10.1093/cid/ciaa794

19. Heavey L, Casey G, Kelly C, Kelly D, McDarby G. No evidence of secondary transmission of COVID-19 from children attending school in Ireland, 2020. Eurosurveillance. 2020;25(21):1–4. doi:10.2807/1560-7917.ES.2020.25.21.2000903

20. Fontanet A, Grant R, Tondeur L, et al. SARS-CoV-2 infection in primary schools in northern France: A retrospective cohort study in an area of high transmission. medRxiv. 2020. doi:10.1101/2020.06.25.20140178

21. Brown NE, Bryant-Genevier J, Bandy U, et al. Antibody responses after classroom exposure to teacher with coronavirus disease, March 2020. Emerg Infect Dis. 2020;26(9):2263–2265. doi:10.3201/eid2609.201802

22. Hildenwall H, Luthander J, Rhedin S, et al. Paediatric COVID-19 admissions in a region with open schools during the two first months of the pandemic. Acta Paediatr Int J Paediatr. 2020;109(10):2152–2154. doi:10.1111/apa.15432

23. National Centre for Immunisation Research and Surveillance. COVID-19 in Schools and Early Childhood Education and Care Services – the Term 3 Experience in NSW.; 2020. https://ncirs.org.au/sites/default/files/2020-10/COVID-1BQZKqdp2CV3QV5nUEsqSg1ygegLmqRygjlsettingsinNSWTerm3report_0.pdf.

24. Ontario Agency for Health Protection and Promotion (Public Health Ontario). COVID-19 Pandemic School Closure and Reopening Impacts. Toronto; 2020.

25. Forbes H, Morton CE, Bacon S, et al. Association between living with children and outcomes from COVID-19: an OpenSAFELY cohort study of 12 million adults in England. medRxiv. 2020:2020.11.01.20222315. https://doi.org/10.1101/2020.11.01.20222315.

26. Ismai, Sharif; Saliba, Vanessa, Lopez Bernal, Jamie; Ramsay, Mary; Ladhani S. SARS-CoV-2 Infection and Transmission in Educational Settings: Cross-Sectional Analysis of Clusters and Outbreaks in England. London; 2020. https://assets.publishing.service.gov.uk/government/uploads/system/uploads/attachment_data/file/911267/School_Outbreaks_Analysis.pdf.

27. Bi Q, Wu Y, Mei S, et al. Epidemiology and transmission of COVID-19 in 391 cases and 1286 of their close contacts in Shenzhen, China: a retrospective cohort study. Lancet Infect Dis. 2020;20(8):911–919. doi:10.1016/S1473-3099(20)30287-5

28. Gostic, Katelyn;Gomez, Ana CR; Mumma Riley O; Kucharski Adam J; Llloyd-Smith JO. Estimated effectiveness of symptom and risk screening to prevent the spread of COVID-19. Elife. 2020;2020;9:e55.

29. Kucharski AJ, Klepac P, Conlan AJK, et al. Effectiveness of isolation, testing, contact tracing, and physical distancing on reducing transmission of SARS-CoV-2 in different settings: a mathematical modelling study. Lancet Infect Dis. 2020;20(10):1151–1160. doi:10.1016/S1473-3099(20)30457-6

30. Voinsky, Irena; Baristaite, Gabriele; Gurwitz D. Effects of age and sex on recovery from COVID-19: Analysis of 5769 Israeli patients. J Infect. 2020;81:e102–e103.

31. Li W, Zhang B, Lu J, et al. The characteristics of household transmission of COVID-19. Clin Infect Dis. 2020;(Xx Xxxx):6–9. doi:10.1093/cid/ciaa450

32. Tuite AR, Fisman DN, Greer AL. Mathematical modelling of COVID-19 transmission and mitigation strategies in the population of Ontario, Canada. Cmaj. 2020;192(19):E497–E505. doi:10.1503/cmaj.200476

33. Chu DK, Akl EA, Duda S, et al. Physical distancing, face masks, and eye protection to prevent person-to-person transmission of SARS-CoV-2 and COVID-19: a systematic review and meta-analysis. Lancet. 2020;395(10242):1973–1987. doi:10.1016/S0140-6736(20)31142-9

34. Wang, S; Kustra, R; Watts, A; Khan K; Mishra S. GTA Proxy Measures of Physical Distancing.; 2020.

35. Cheng HY, Jian SW, Liu DP, Ng TC, Huang WT, Lin HH. Contact Tracing Assessment of COVID-19 Transmission Dynamics in Taiwan and Risk at Different Exposure Periods before and after Symptom Onset. JAMA Intern Med. 2020;180(9):1156–1163. doi:10.1001/jamainternmed.2020.2020

