## Supplemental appendix, tables and figures for "The potential impact of School Closure Relative to Community-based Non-pharmaceutical Interventions on COVID-19 Cases in Ontario, Canada"

### eAppendix

David Naimark, Sharmistha Mishra, Kali Barret, Yasin A. Khan, Stephen Mac, Raphael Ximenes, Beate Sander

**Table of Contents**

**eMethods**……………………………………………………………………………………………………...pg 3

**eTable 1.** Logistic coefficients for imputation of missing symptomatic status and for assignment of symptomatic status of individuals infected with COVID-19 in the Agent Based Model of COVID-19 Transmission (ABMCT)……………………………………………………………………………………………..pg 7

**eTable 2.** Percentage of essential workers by Social Policy Simulation Database and Model (SPSD/M) industry designation……………………………………………………………………………………………pg 8

**eTable 3.** Summary of simulation scenarios……………………………………………………………......pg 9

**eTable 4.** Model simulation output for classroom closures and location of acquisition of infection……………..pg 10

**eFigure 1.** Agent-based Model of COVID-19 Transmission (ABMCT) model schematic………………………pg 11

**eFigure 2**. Demonstration of 5000 samples from the multivariate lognormal distribution of daily contacts at work………………………………………………………………………………………………………pg 12

**eFigure 3**. Demonstration of 5000 samples from the multivariate lognormal distribution of potential numbers of daily contacts at daycare, and primary, elementary, and high schools……………………………………………pg 13

**eFigure 4**. Demonstration of the correlation of 5000 samples from the multivariate lognormal distribution of potential numbers of daily contacts at work and within the urban city or rural region……………………………………………………………………………………………………...pg 14

**eFigure 5**. Observed versus smoothed daily, new, symptomatic, COVID-19 case counts – first wave – for various Gaussian kernel smoothing bandwidths…………………………………………………………………………...pg 15

**eFigure 6**. Observed versus smoothed daily, new, symptomatic, COVID-19 case counts – first wave – for the selected Gaussian kernel smoothing bandwidth of 7 days………………………………………………………...pg 16

**eFigure 7**. Model calibration to daily new, symptomatic, confirmed COVID-19 cases in Ontario – first wave……………………………………………………………………………………………………pg 17

**eFigure 8**. Observed versus smoothed daily, new, symptomatic, COVID-19 case counts – second wave – for various Gaussian kernel smoothing bandwidths…………………………………………………………………….....pg 18

**eFigure 9**. Observed versus smoothed daily, new, symptomatic, COVID-19 case counts – second wave – for the selected Gaussian kernel smoothing bandwidth of 2 days………………………………………………………...pg 19

**eFigure 10**. Model calibration to daily new, total, confirmed COVID-19 cases in Ontario – second wave……..pg 20

**eFigure 11**. Observed versus smoothed daily, new, symptomatic, COVID-19 case counts – during the first 15 days of October, 2020 – for various Gaussian kernel smoothing bandwidths…………………………………………pg 21

**eFigure 12**. Observed versus smoothed daily, new, symptomatic, COVID-19 case counts – during the first 15 days of October, 2020 – for the selected Gaussian kernel smoothing bandwidth of 2 days……………………………pg 22

**eFigure 13**. Model calibration to daily new, total, confirmed COVID-19 cases in Ontario – first 15 days of October, 2020……………………………………………………………………………………………………..pg 23

**eFigure 14**. Distribution of numbers of daycare, primary, elementary and high school classroom closures for scenarios in which schools had re-opened……………………………………………………………………..pg 24

**eFigure 15**. Distribution of the percentage of COVID-19 infections acquired in schools for students and teachers in scenarios in which schools had re-opened……………………………………………………………………..pg 25

**eFigure 16**. Model simulation results for scenario 1a……………………………………………………………pg 26

**eFigure 17**. Model simulation results for scenario 1b……………………………………………………………pg 27

**eFigure 18**. Model simulation results for scenario 2a……………………………………………………………pg 28

**eFigure 19**. Model simulation results for scenario 2b……………………………………………………………pg 29

**eFigure 20**. Model simulation results for scenario 3a……………………………………………………………pg 30

**eFigure 21**. Model simulation results for scenario 3b……………………………………………………………pg 31

**eFigure 22**. Reduction of cumulative cases between September 1 and October 31, 2020, attributable to the two policy choices to implement public health restrictions or not vs. to open schools or not………………………....pg 32

**eReferences**………………………………………………………………………………………pg 33

**eMethods**

**Development of a Synthetic Population Representative of Ontario, Canada**

We developed a synthetic population of one million individuals representative of the inhabitants of Ontario, Canada (population ~ 14.5 million) based on data within the Social Policy Simulation Database and Model (SPSD/M) [1] developed by Statistics Canada. SPSD/M contains data on households in Ontario categorized by rural or urban location, numbers of household members, the age and sex of household members, labour force participation and industry. Household types consist of the 39,774 combinations of the latter factors. Each type has an associated household weight representing the number of households in Ontario with that particular configuration. For the synthetic population, 413,957 household were randomly sampled from the available household types, according to the household weights, to yield 1 million hypothetical individuals.

Households were randomly assigned to 52 cities and one rural region according to the SPSD/M urban/rural designation of the associated household type. The 2020 populations of these regions were extrapolated from 2011 and 2016 census data to 2020 and then reduced to model scale. Within urban settings, households were further randomly assigned to neighborhoods which were configured as square tiles with areas of 4 $km^2$ such that the number of individuals in a neighborhood was determined by the population density of the city. A similar process assigned rural household to districts that had an area of 64 $km^2$ and a population density equal to the average for rural Canada of 150/$km^2$ [2].

Children less than 2 years old were assumed to remain within their household or accompany their parents into neighborhood/districts and regions. Older children spent time within the household, neighborhood/district and region but also could potentially go to school on weekdays during the school year (subject to general school opening or closure and specific restrictions, see the main text for details). Children between 2 and 3 years old were assigned to daycare settings with a cap of 10 children per daycare, children between 4 and 13 were assigned to primary/elementary schools with a classroom cap of 23 students and children between 14 and 17 were assigned to high schools (secondary schools) with a classroom cap of 15. Daycare centres were assumed to draw children from a single neighborhood or district. On model scale, primary/elementary and high school enrollments were limited to 150 students. These schools drew students from multiple contiguous neighborhoods or districts. Each daycare or classroom was assigned a teacher who was randomly sampled from adults in the region whose SPSD/M industry designation was 11, educational services.

Young adults, 18 to 34 years old, could join the workforce or potentially go to college or university (a post-secondary institution). The probability of the latter was based on the total enrollment among colleges and universities in Ontario divided by the size of the 18 to 34 year-old population in the province. The attendance of a student at a given institution was selected randomly according to the proportion of post-secondary students in Ontario enrolled at that college or university. Colleges and universities were assumed to draw students from all regions (e.g. a student whose household was located in the city of Barrie, Ontario, could attend the University of Toronto located in Toronto). When post-secondary institutions were in session, students could spend time on campus or in the region in which the institution is located. During the summer break or during general college or university closures (which was assumed to be the case after March 15, 2020), students returned to their households and spent time in the associated neighborhood/district and region.

Adults, 18 to 34 yeas old, not enrolled in a post-secondary institution, and 35 to 64 years old, could be members of the workforce. Workplaces were characterized by industry type, region, and workplace size. Workplace sizes were categorized as extremely small (1 – 10 workers), small (11 – 99 workers), medium (100 – 499 workers), and large (> 500 workers). Initially, the number of potential workplace assignments in each combination of these three factors was obtained by taking the product of the three marginal proportions and the synthetic population size of 1 million individuals. Then, because some combinations of industry and region yielded too few potential workplace assignments for medium and large workplaces, they were re-assigned to larger population centres. Working age adults were then randomly assigned to workplaces in their region according to their SPSD/M industry designator. If a workplace could not be found for an individual within a city or region, an alternate workplace in an adjacent city was sought. If no workplaces could be identified in the back-up city, a placement was sought in the largest population centre, Toronto. If no workplace could be identified, the person was designated as unemployed and could spend time within their household, neighborhood or district, and region. Employed, working-age adults spent time at home, within their neighborhood or district and, on weekdays, at work.

Adults, 65 and older, were assumed to be retired from the workforce and could spend time in their households, neighborhoods or districts, and regions.

**ABMCT Model Structure**

The Agent-based Model for COVID-19 Transmission (ABMCT) was structured to model the COVID-19 pandemic in Ontario and made use of the synthetic population. Each member of the synthetic population was modelled as an agent that could interact with other agents and potentially transmit COVID-19 in the settings described above.

For the first wave, the model was seeded with 150 infectious individuals who were selected from the synthetic population at random on February 22, 2020. Each of the seed individual's number of days of prior infectiousness was drawn from a uniform distribution. There was a seven-day run-in period until February 29, 2020 (day 0) when cases began to be reported by the model.

A schematic of the transmission model is shown in **eFigure 1** and the input parameters in **Table 1** in the main text.

Except for the persons seeding the model, all synthetic individuals were initially susceptible to COVID-19. They could contact and be exposed to infectious individuals within their household, at school, at college or university, at workplaces, in neighborhoods or in regions. Exposed individuals may not have been infected and remain susceptible or they may have become infected. Infected individuals were unable to transmit the virus during a four-day latent period, after which, infected individuals became infectious and able to transmit the virus. Infectious individuals may or may not have developed symptoms. The former entered a one-day pre-symptomatic stage (i.e. a five-day incubation period) followed by a symptomatic stage. The duration of infectiousness for both individuals with and without symptoms was modelled to be 15 days.

The proportion of symptomatic COVID-19 cases by ten-year age category was obtained from Ontario's Case and Contact Management (CCMPLUS) database.[3] In order to calculate these proportions, missing symptomatic status in the CCMPLUS data was imputed by constructing a logistic regression equation employing data from individuals with known symptomatic status and with sex and age category as covariates (**eTable1**). The symptomatic status of an individual, i, with missing status in the database, was calculated as:

$$\ln(odds_i) = \beta_0 + \beta_1 X_{1i} + \cdots + \beta_n X_{ni}$$

$$odds_i = exp(\beta_0 + \beta_1 X_{1i} + \cdots + \beta_n X_{ni})$$

$$P_i = {odds_i}/{(1 + odds_i)}$$

$$S_i \sim Bernoulli(P_i).$$

That is, the binary indicators for the person's age category and sex allowed calculation of the odds of being symptomatic which, in turn, allowed calculation the person's probability of being symptomatic, $P_i$. The individual's binary symptomatic status, $S_i = 1$ for symptomatic and $S_i = 0$ for non-symptomatic, was drawn from a Bernoulli distribution with a mean of $P_i$.

A similar method was used in the ABMCT model to assign symptomatic status to individuals infected with COVID-19 given the age and sex designation for an individual sampled along with that person's other characteristics from SPSD/M [1]. Since, the SPSD/M data source contains only male and female sex, the logistic regression equation was modified to fold the 'Other sex' effect into a revised intercept value:

$$\ln(odds) = \beta_0 + \beta_1 \bar{X}_1 + \cdots + \beta_n \bar{X}_n$$

$$\beta_0 = \ln(odds) - \beta_1 \bar{X}_1 - \cdots - \beta_n \bar{X}_n$$

$$\beta_0' = \beta_0 + \beta_n \bar{X}_n = \ln(odds) - \beta_1 \bar{X}_1 - \cdots - \beta_{n-1} \bar{X}_{n-1}$$

$$\ln(odds) = \beta_0' + \beta_1 \bar{X}_1 + \cdots + \beta_{n-1} \bar{X}_{n-1} ,$$

where $\bar{X}_n$ is the average of the covariate for 'Other sex' which, since the covariate is a categorical binary, equals the proportion of the CCMPLUS cohort with 'Other' as the listed sex.

Symptomatic individuals could present to an emergency department (ED) where they could either be admitted to hospital or sent home to isolate until recovered. Symptomatic individuals who did not visit an ED could self-isolate until recovery. A proportion of symptomatic cases would present for testing and be recorded as confirmed cases while a proportion of asymptomatic cases could be detected through contact tracing, reported as confirmed cases, and quarantined until recovery. The probability of detection, i.e. of an infectious individual being a confirmed case, during the first wave of COVID-19 was estimated to be 20% by Tuite et al[4]. in their compartmental transmission model. In addition, we estimated the probability of detection of an infectious individual in Ontario separately, for symptomatic and asymptomatic cases, via the cumulative number of confirmed cases until June 9, 2020, and the seroprevalence of anti-Covid19 antibodies reported by Public Health Ontario until that date of ~1%.[5] As a simplifying assumption, the proportions of symptomatic and asymptomatic cases reported in the CCMPLUS database in June, 2020, 0.79 and 0.21, respectively, were applied to the general population in the calculation of detection probabilities. For symptomatic individuals:

pDetect|symptomatic ~ cumulative cases/(0.79*population of Ontario*0.01) = 22394/(0.79*14.5M*0.01) = 0.195,

and for asymptomatic individuals:

pDetect|asymptomatic ~ 4244/(0.21*14.5M*0.01) = 0.139.

Given the uncertainty in these estimates, the probabilities of detection were refined via model calibration as described below.

The absorbing states in the ABMCT were 'recovered' or 'admitted to hospital' - where individuals could either die or recover. Death outside of hospital, such as in a long-term care (LTC) facility, was not considered in the current iteration of the model nor was nosocomial transmission within hospitals.

Susceptible persons were modelled to be potentially infected via close contact with an infectious individual. Mixing of individuals was assumed to occur randomly within households, classrooms, college or university campuses, workplaces, neighborhoods or districts, and regions. The average number of close contacts per day, prior to the institution of public health restrictions in March, and April 2020, in the settings mentioned above was based on the CONNECT study [6] (**Table 1** main text). For a given individual, the number of household contacts per day was assumed to be equal to the number of individuals in the household. The number of potential close contacts in non-household settings was obtained by rounding samples from a multivariate lognormal distribution to the nearest integer assuming a standard deviation of the log counts of 20% of the mean log count and correlation coefficients among settings of 0.9. This ensured that potential close contact numbers were always positive, that there was a rightward skew in the distribution of contact numbers such that a small proportion of individuals had very high potential daily contact numbers, and that a sampled daily close contact number in one setting was correlated with the number in other settings. The latter sampling provided the number of potential close contacts. The actual number of close contacts during a day could be constrained by the total number of individuals in a particular setting (in a workplace, classroom, etc.) (**eFigures, 2, 3, 4**).

**Calibration to Ontario's daily, new, symptomatic, confirmed case counts – first wave**

For the first wave of COVID-19 in Ontario we calibrated the daily number of new, confirmed, symptomatic cases outside of LTC facilities reported by the model from February 29 until July 7, 2020, to the observed number of similarly defined cases in the CCMPLUS database.[3] We assumed an initial unmitigated rise in infections from the origin of the model on February 22, 2020, until March 7, 2020, at which time there was a modelled spontaneous reduction in close contact numbers, followed by closure of schools, colleges and universities on March 15, 2020, and, ultimately, closure of workplaces, with the exception of essential workers, which had an effect on April 8, 2020 (**Table 1** in the main text).

The proportion of workers who were essential was estimated for each SPSD/M industry type as one minus the proportion of teleworkers in the particular industry derived from Statistics Canada data obtained on May 29, 2020[7] (**eTable2**). Assignment of an individual worker to essential status was by means of drawing from a Bernoulli distribution with a mean equal to the proportion of essential workers for that individual's SPSD/M industry designator.

Calibration was achieved via a combination of manual search of the calibration parameters and via the bound optimization by quadratic approximation (BOBYQA) directed search algorithm [8] which seeks to minimize a goodness-of-fit statistic defined as the sum of squared differences between modeled and observed daily case counts. To ease the calibration process, observed and modelled case counts were subjected to Gaussian kernel smoothing:

$$\widetilde{c_t} = \frac{\sum_i^N K(t,i) \cdot c_i}{\sum_j^N (t,i)}$$

where a smoothed count, $\widetilde{c_t}$, at a particular time, t, is a weighted average of all of the case counts, $c_i$, from the first day to the Nth day, the last day in the dataset, with a Gaussian, kernel weighting function:

$$K(t,i) = exp\left(-\frac{(t-i)^2}{2b^2}\right)$$

where with 'b', is the bandwidth in days.

For first wave calibration, b = 7 days, provided an optimum of reduced stochasticity and a smoothed peak value close to the un-smoothed peak (**eFigures 5 and 6**).

Final model parameters of the calibrated model are presented in **Table 1** in the main text, and calibrated model results for the number of daily, new, symptomatic, confirmed cases in Ontario from February 29 to July 7, 2020, are presented in e**Figure 7**.

**Calibration to Ontario's daily new, confirmed case counts in September, 2020 – second wave**

For the COVID-19 second wave in Ontario, we obtained the cumulative number of confirmed cases, symptomatic and asymptomatic by ten-year age category from the CCMPLUS database. These counts were the inflated by dividing them by the case detection probabilities described above according to symptomatic status. The sum of these two counts was reduced to model scale and that number of individuals within the synthetic population were selected at random within the ten-year age categories to represent recovered and immune or deceased individuals as a result of COVID-19 infection in the first wave. The model was reseeded with randomly selected new infectious individuals, the number of whom was determined through model calibration. Contact numbers in settings outside households were adjusted to be lower than the pre-pandemic values according to the proportion of time that mobile telephones were outside of households in major Ontario cities in August, 2020, compared to the values prior to the pandemic.[9] After a run-in period from August 15 to 31, the model began reporting daily, new, confirmed, cases of COVID-19 on September 1, 2020. For calibration, the indicator for school re-opening was turned off, i.e. modelled schools remained closed, under the assumption that an effect of school opening in the observed case counts would take longer than two weeks to manifest. For Gaussian smoothing of observed daily case counts, a bandwidth of 2 days fit the data the best (**eFigures 8 and 9**). Using methods described above, we re-calibrated the model to the number of cases between September 1 and 30, 2020 (**eFigure 10**).

**Calibration to Ontario's daily new, confirmed case counts from October 1 – 15, 2020**

For the first two weeks of October, there was an observed decrease in the rate of growth of new, confirmed COVID-19 cases to 0.8% per day in the CCMPLUS database. We replicated this reduction in growth by allowing modelled transmissions to occur randomly but limiting their number to 0.8% of the existing infectious individuals per day. For calibration, the indicator for school re-opening was turned on, i.e. modelled schools re-opened, under the assumption that an effect of school opening in mid-September would be reflected in the observed case counts would in early October, 2020. Using methods described above, we re-calibrated the model to the number of cases between September 30 and October 15, 2020 (**eFigure 11**).

| Factor | Beta coefficient | SE | p-value |
|---|---|---|---|
| Intercept | -0.70182 | 0.075148 | <0.001 |
| Age group 10 to 19 | -0.47205 | 0.091769 | <0.001 |
| Age group 20 to 29 | -0.76417 | 0.080276 | <0.001 |
| Age group 30 to 39 | -0.86852 | 0.081766 | <0.001 |
| Age group 40 to 49 | -0.92607 | 0.082403 | <0.001 |
| Age group 50 to 59 | -1.02056 | 0.082055 | <0.001 |
| Age group 60 to 69 | -0.77826 | 0.084047 | <0.001 |
| Age group 70 to 79 | -0.60985 | 0.088948 | <0.001 |
| Age group 80 or older | -0.10229 | 0.080993 | 0.207 |
| Male sex | 0.067427 | 0.026434 | 0.011 |
| Other sex[a] | 1.309702 | 0.145335 | <0.001 |

**eTable 1. Logistic coefficients for imputation of missing symptomatic status and for assignment of symptomatic status of individuals infected with COVID-19 in the Agent Based Model of COVID-19 Transmission (ABMCT)** - data on symptomatic status was obtained from the Case and Contact Management (CCMPLUS) database for COVID-19 cases in Ontario.[3] For individuals with missing symptomatic status, we developed a logistic regression model, with coefficients listed in the table in order to perform imputation as described in the eMethods. SE – standard errors of the logistic beta coefficients.

[a] 'Other sex' refers to individuals who neither identify as male or female.

| SPSD/M Industry Type | SPSD/M industry designation | Percentage of essential workers |
|---|---|---|
| Not applicable | 0 | 43.2% |
| Agriculture | 1 | 61.4% |
| Other primary | 2 | 43.2% |
| Utilities | 3 | 43.2% |
| Construction | 4 | 60.9% |
| Manufacturing | 5 | 54.6% |
| Trade | 6 | 50.9% |
| Transportation and warehousing | 7 | 42.7% |
| Finance, insurance, real estate, and leasing | 8 | 35.9% |
| Professional, scientific, and technical services | 9 | 14.1% |
| Management, administration, and other support | 10 | 49.8% |
| Educational services | 11 | 33.7% |
| Health care and social assistance | 12 | 41.1% |
| Information, culture, and recreation | 13 | 31.4% |
| Accommodation and food services | 14 | 65.1% |
| Other services | 15 | 45.2% |

**eTable 2. Percentage of essential workers by Social Policy Simulation Database and Model (SPSD/M) industry designation** – the proportion of workers who were essential was estimated for each SPSD/M industry type as one minus the proportion of teleworkers in the particular industry derived from Statistics Canada data obtained on May 29, 2020[7]. For ABMCT sampling purposes, the percentages were converted to proportions so that an individual simulated worker's essential status could be sampled from a Bernoulli distribution as described in the eMethods.

| | | Nonpharmaceutical Intervention Scenarios | | |
|---|---|---|---|---|
| | | 1: No restrictions imposed on October 1, 2020 | 2: Restrictions imposed on October 1, 2020 | 3: Replication of case counts, October 1- 15, 2020 |
| School Re-opening Scenarios | A. Schools remained closed | 1A | 2A | 3A |
| | B. Schools re-opened on September 15, 2020 | 1B | 2B | 3B |

**eTable 3. Summary of simulation scenarios –** Schools remained closed – daycare facilities and primary, elementary and high schools remained closed on Sept 15, 2020; Schools re-opened on Sept 15, 2020: daycare facilities and primary, elementary and high schools re-opened on that date with safety measures as indicated in the text; no restrictions: the rise in daily new, confirmed, COVID-19 case counts observed between September 1 – 30, 2020, persisted until October 31, 2020; restrictions – on October 1, 2020, workplaces were closed except for essential workers, the population remained at home except for essential outings thereby reducing non-household contacts and probability of transmission of COVID-19 between contacts by 50% compared to the August, 2020, baseline; replication of case counts: a rise in daily new, confirmed, COVID-19 case counts observed between September 1 – 30, 2020, followed by a reduction in the rate of new case counts to 0.8% per day as observed in the first two weeks of October, 2020. For all public health scenarios, simulation data was reported by the model from September 1 to October 31, 2020.

| | No restrictions imposed, median (IQR) | Restrictions imposed, median (IQR) | Replicating observed counts, median (IQR) |
|---|---|---|---|
| **Number of any classroom/daycare closures** | 1726 (421) | 1051 (337) | 587 (178) |
| **Number of daycare closures** | 87 (44) | 44 (44) | 15 (15) |
| **Number of primary/elementary classroom closures** | 1175 (279) | 725 (236) | 406 (116) |
| **Number of high school classroom closures** | 479 (178) | 276 (91) | 160 (73) |
| **Number of COVID-19 infections acquired in school** | 493 (134) | 421 (109) | 167 (58) |
| **Number acquired outside of school** | 15892 (1842) | 9643 (1407) | 7265 (848) |
| **Percentage of COVID-19 acquired in school** | 3.15% (0.76%) | 4.19% (0.80%) | 2.37% (0.84%) |

**eTable 4. Model simulation output for classroom closures and location of acquisition of infection -** numbers of confirmed infections correspond to those among children in daycare (ages 2-3), primary and elementary school (ages 4- 13), or high school (ages 14 – 17) from September 1 to October 31, 2020. Infections and classroom closure numbers are on the model scale of 1 million individuals rather than the Provincial scale of 14.5 million individuals. IQR – interquartile range.

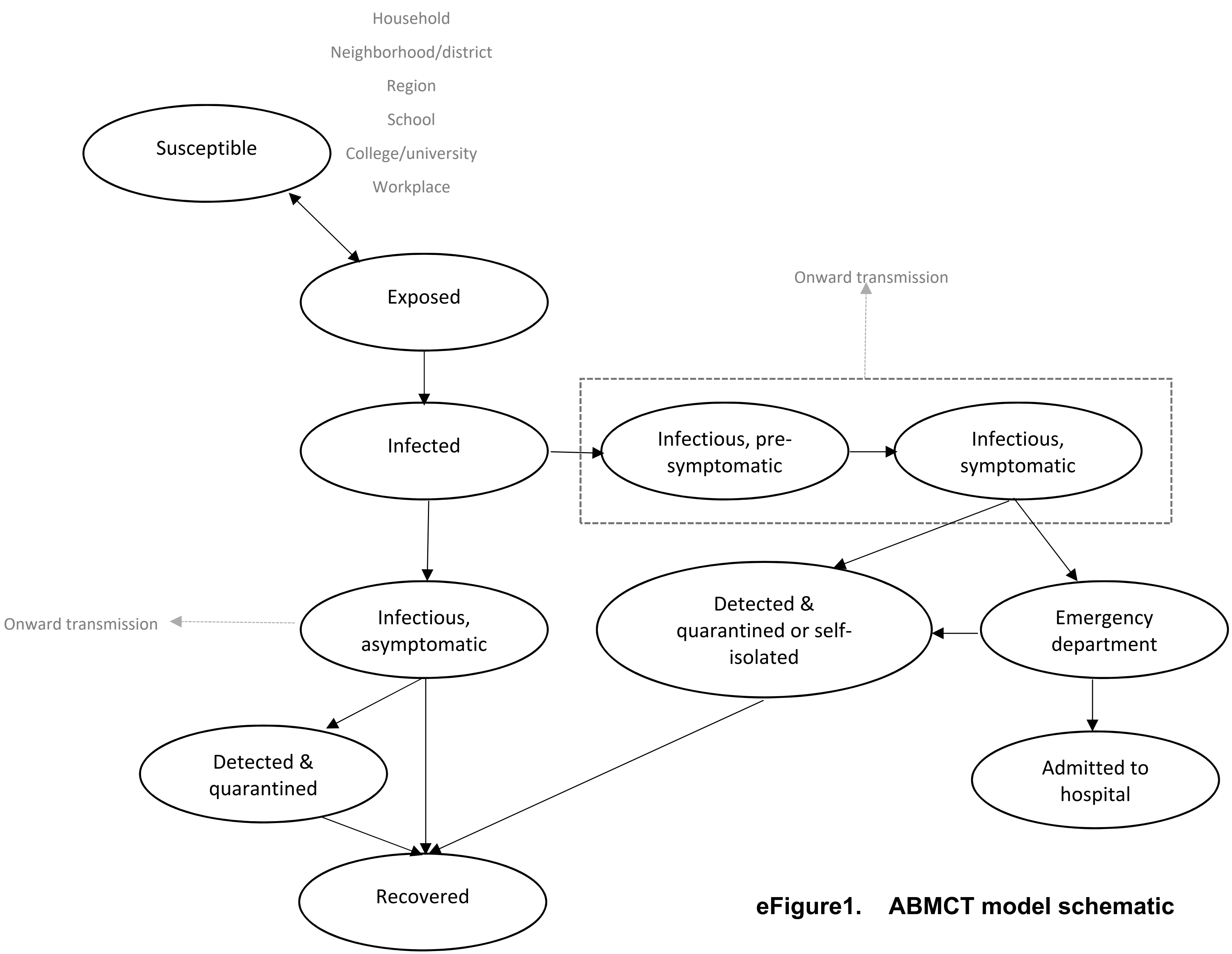


**eFigure1. ABMCT model schematic**

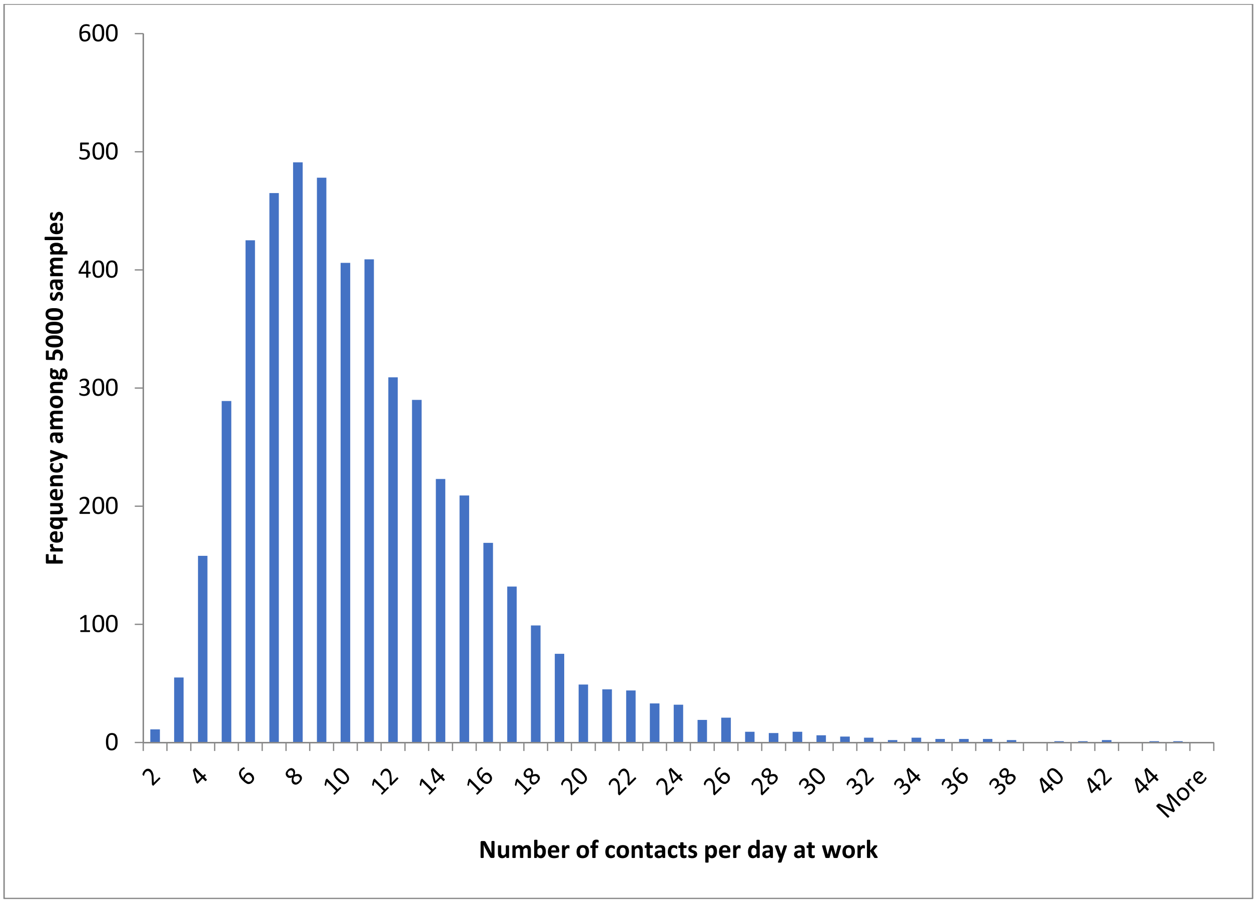


**eFigure 2. Demonstration of 5000 samples from the multivariate lognormal distribution of daily contacts at work.** An individual sample produced the natural logarithm of work contacts which was then exponentiated and rounded to the nearest integer. The actual number of daily contacts may be lower than the sampled number of potential daily contacts in smaller workplaces with fewer numbers of workers.

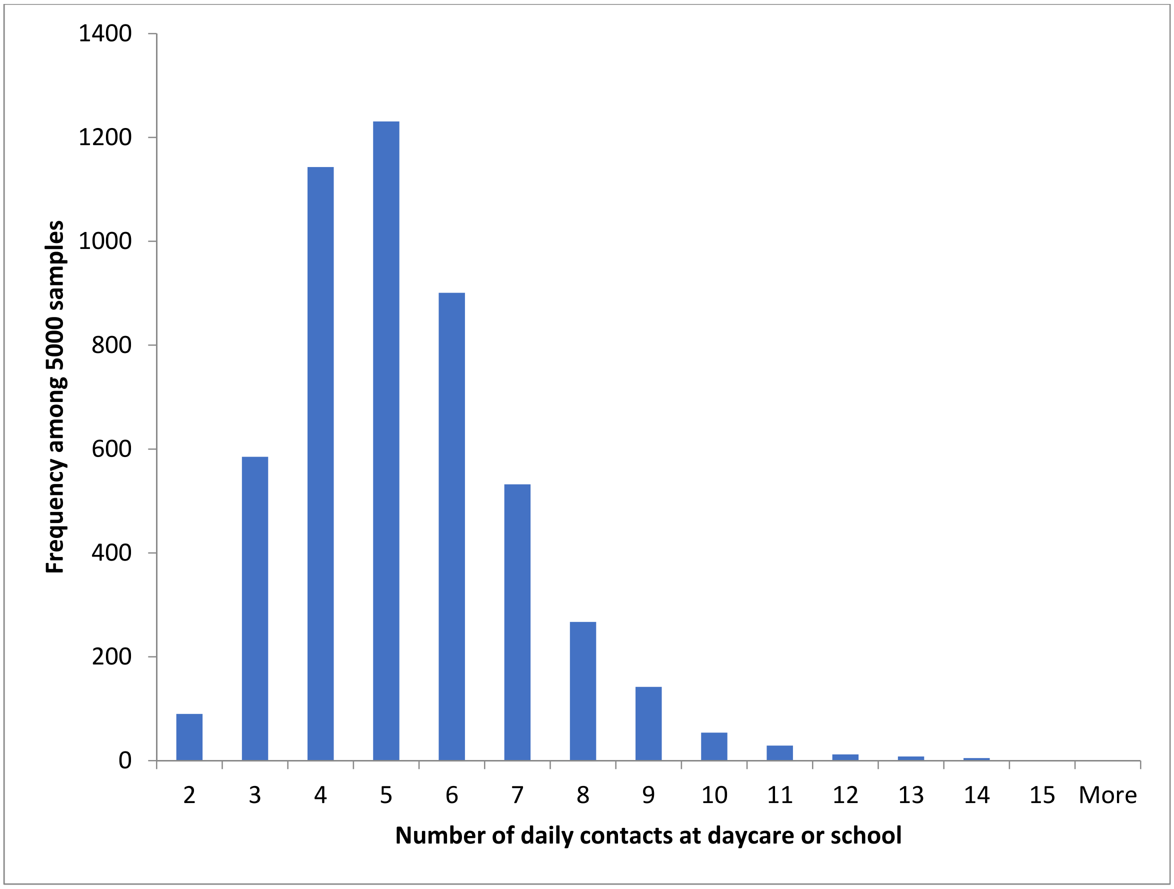


**eFigure 3. Demonstration of 5000 samples from the multivariate lognormal distribution of potential numbers of daily contacts at daycare, and primary, elementary, and high schools.** An individual sample produced the natural logarithm of contacts which was then exponentiated and rounded to the nearest integer. The actual number of daily contacts may be lower than the sampled number of potential daily contacts because of caps on daycare facility and schoolroom capacity.

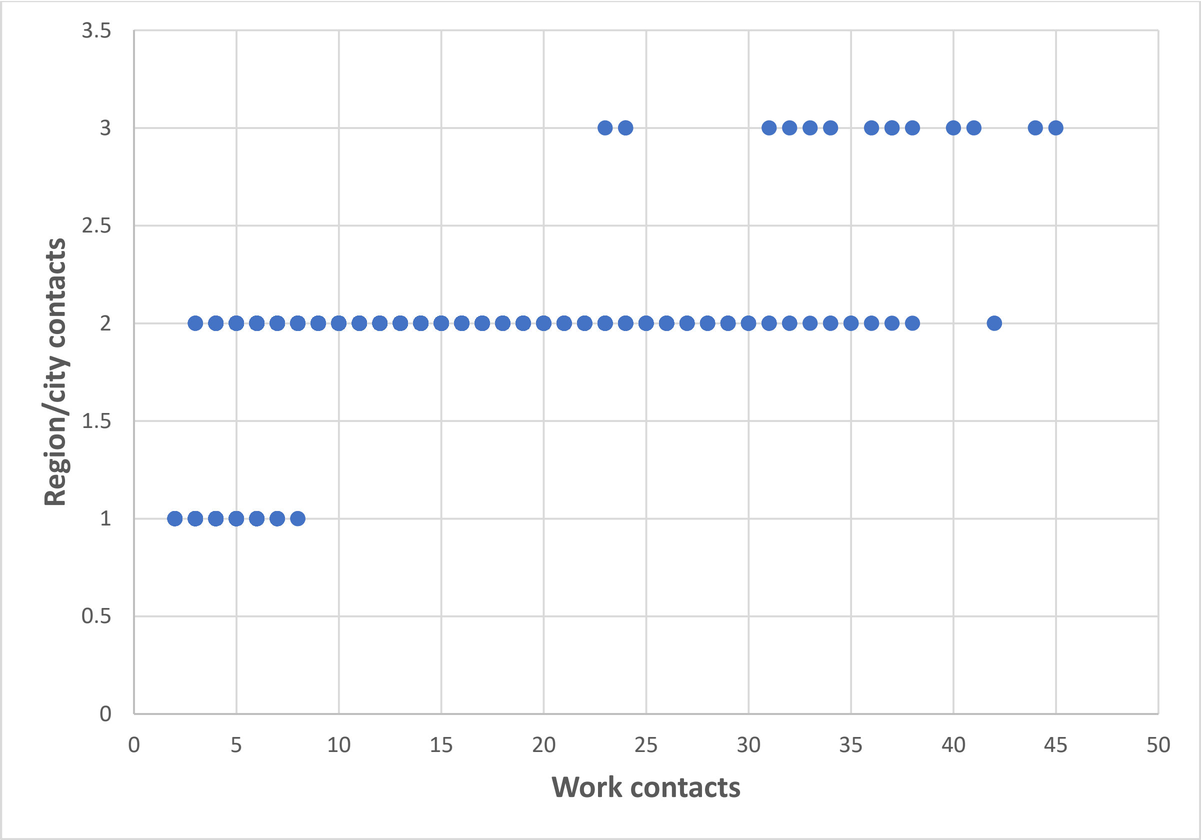


**eFigure 4. Demonstration of the correlation of 5000 samples from the multivariate lognormal distribution of potential numbers of daily contacts at work and within the urban city or rural region.** Note that each point on the graph represents more than one sample.

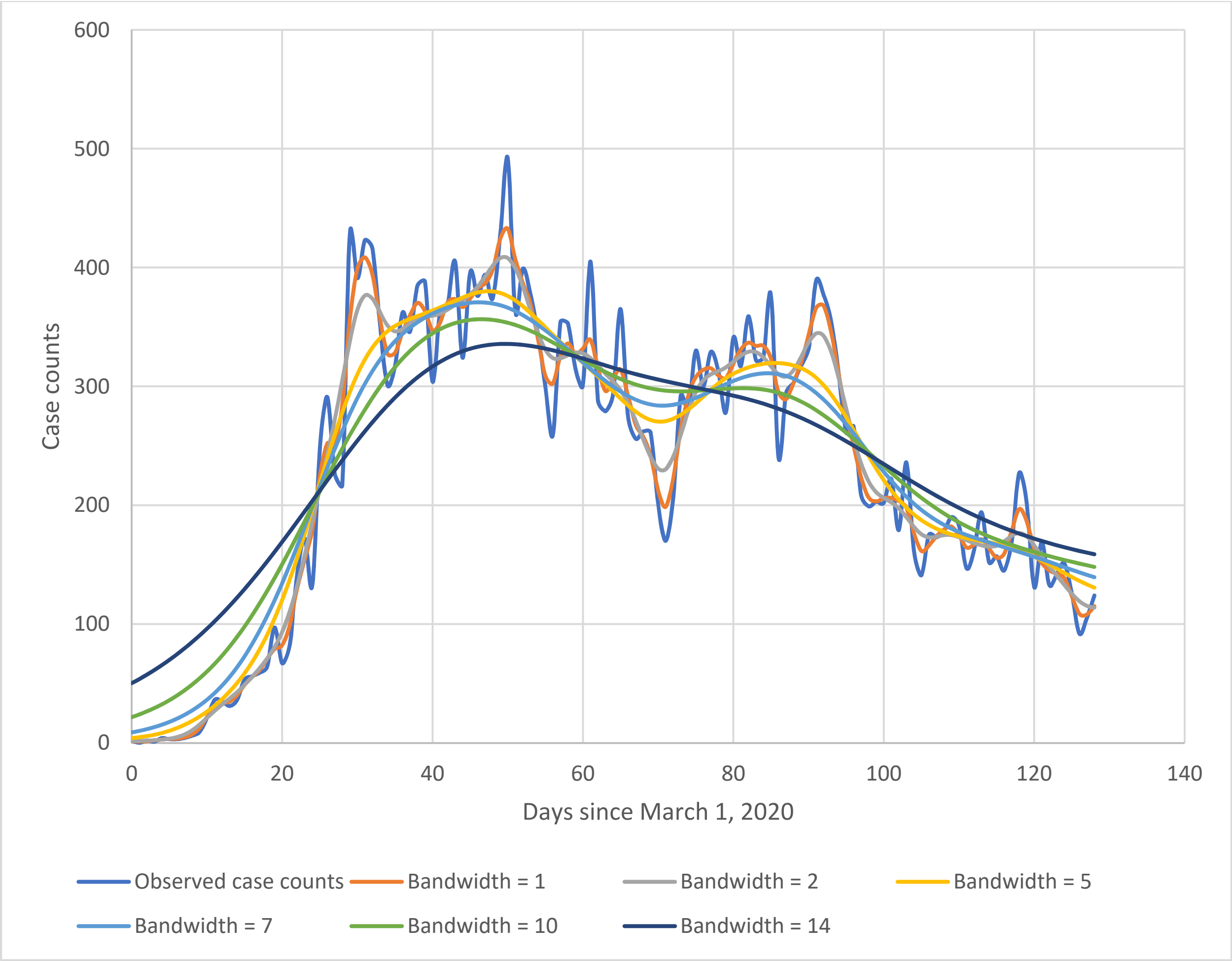


**eFigure 5. Observed versus smoothed daily, new, symptomatic, COVID-19 case counts – first wave – for various Gaussian kernel smoothing bandwidths.**

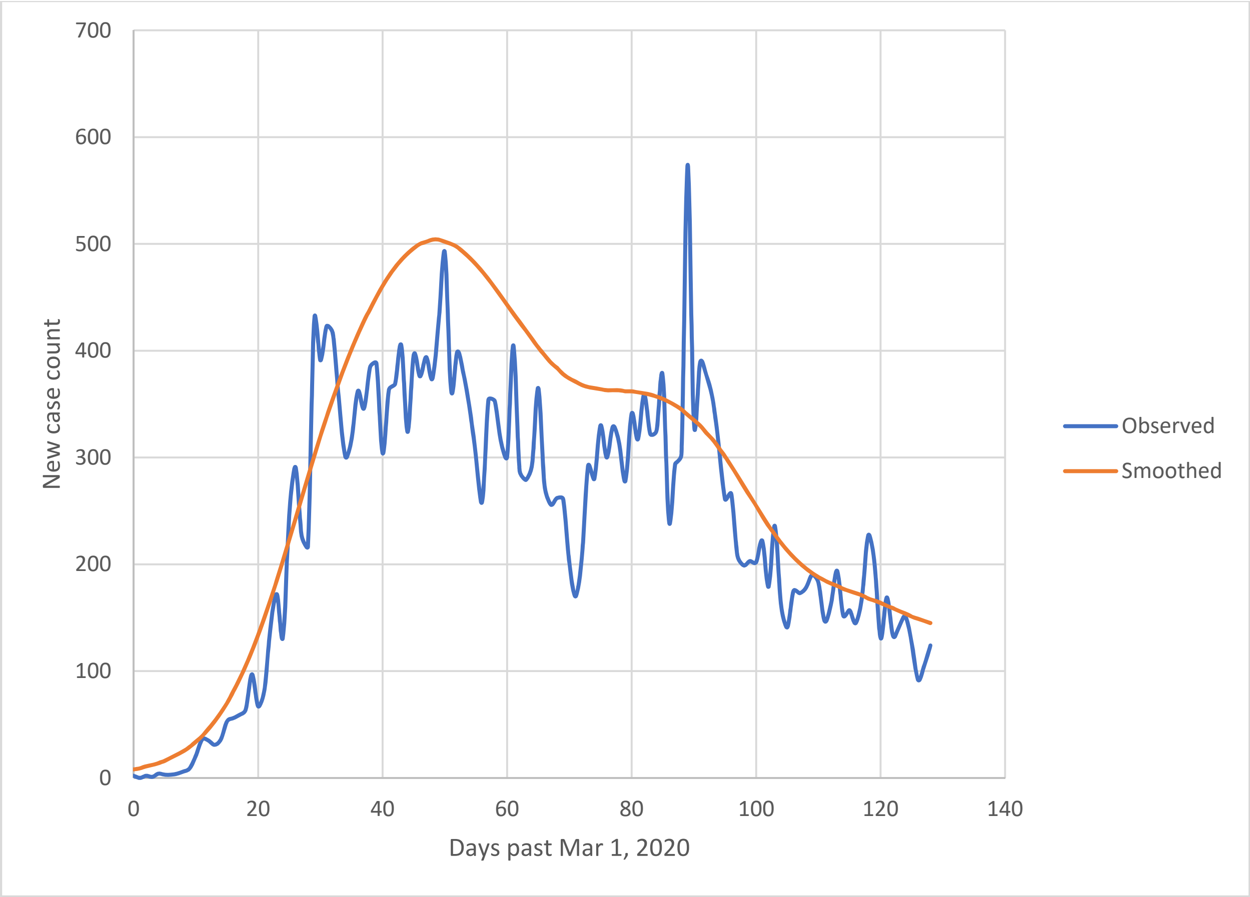


**eFigure 6. Observed versus smoothed daily, new, symptomatic, COVID-19 case counts – first wave – for the selected Gaussian kernel smoothing bandwidth of 7 days.**

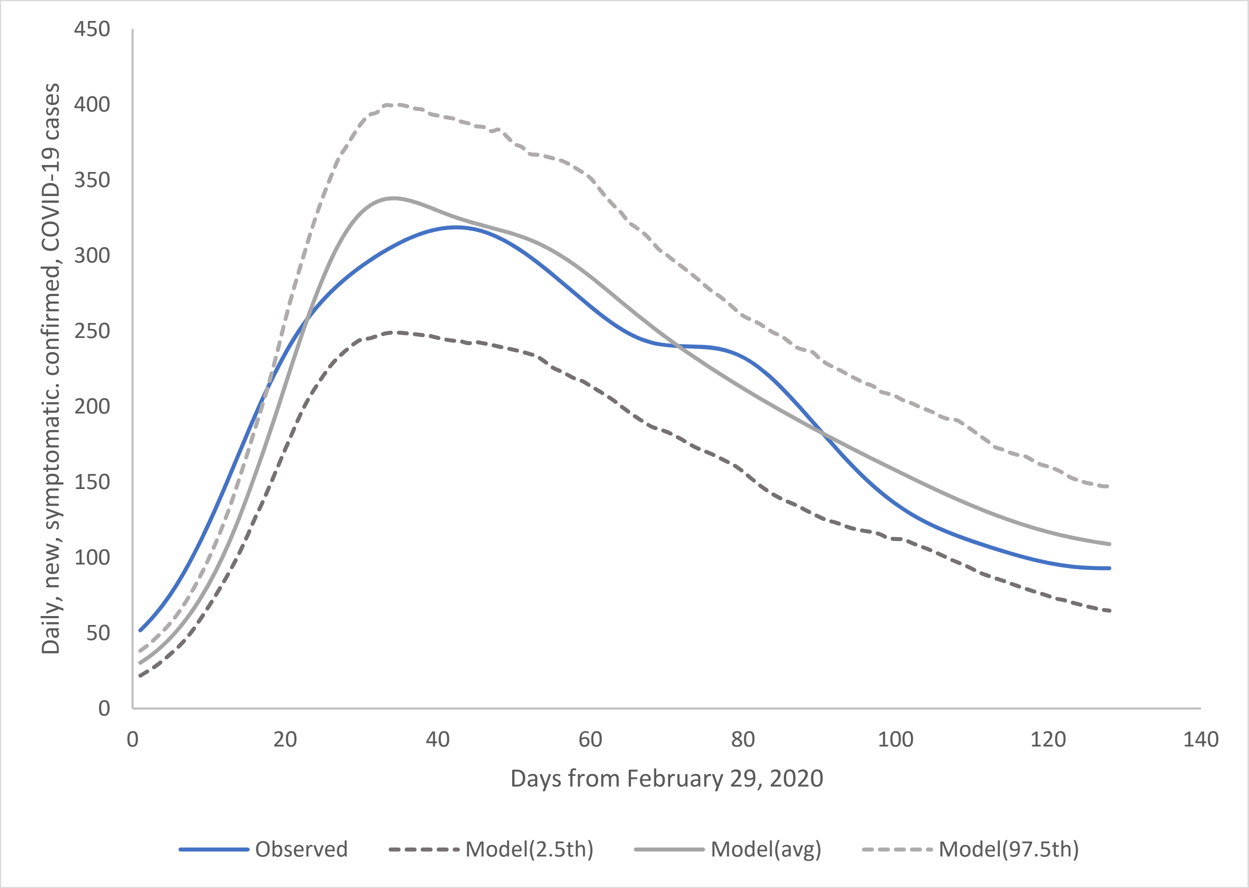


**eFigure 7. Model calibration to daily new, symptomatic, confirmed COVID-19 cases in Ontario – first wave.** Observed cases in the CCMPLUS database (blue) compared to the average among 250 model replicates (solid grey) along with the associated 2.5$^{th}$ and 97.5$^{th}$ percentiles (dotted grey) between February 29 (day 0) and July 7, 2020. Note, both observed and modelled counts were subjected to Gaussian kernel smoothing with a bandwidth of 7 days.

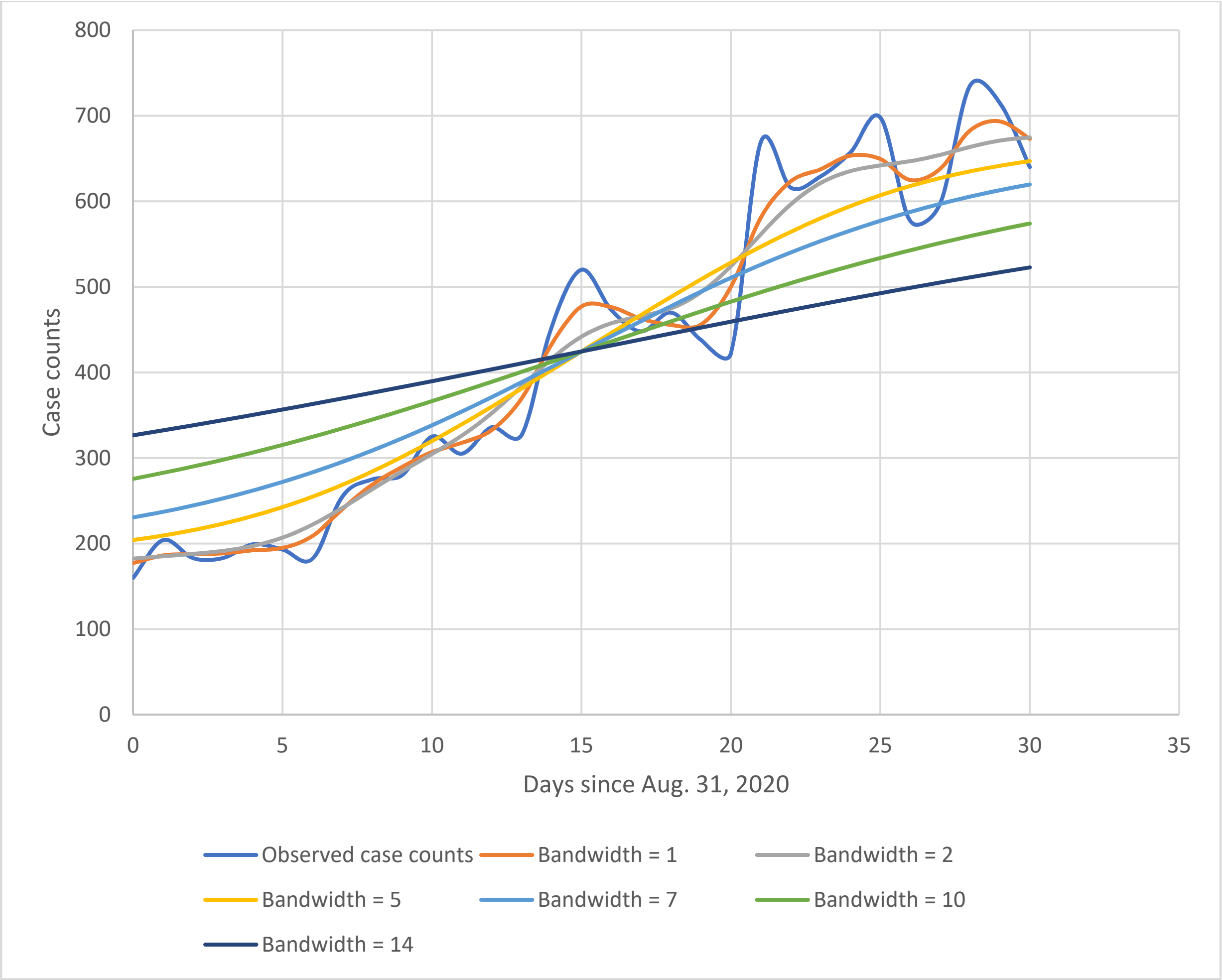


**eFigure 8. Observed versus smoothed daily, new, symptomatic, COVID-19 case counts – second wave – for various Gaussian kernel smoothing bandwidths.**

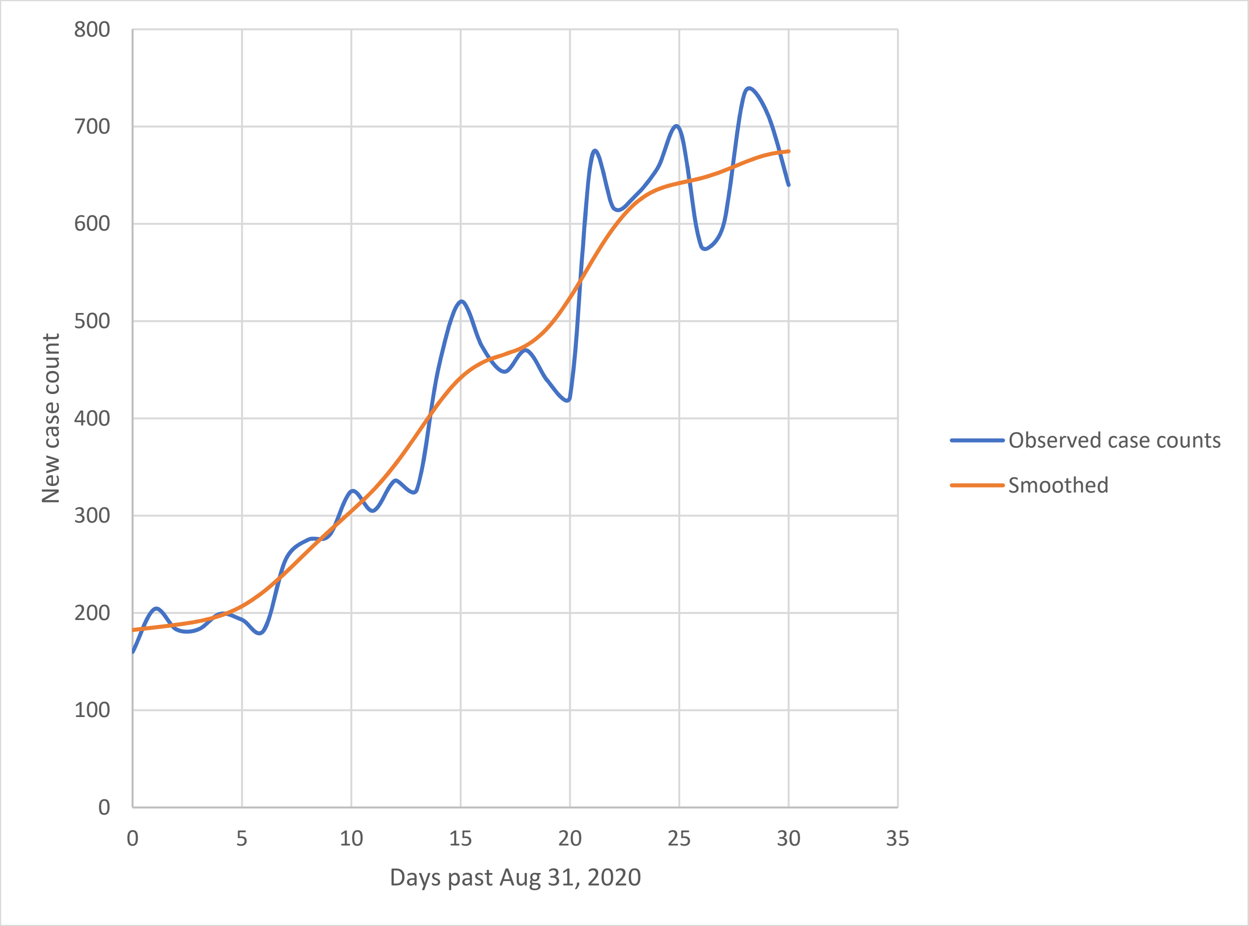


**eFigure 9. Observed versus smoothed daily, new, symptomatic, COVID-19 case counts – second wave – for the selected Gaussian kernel smoothing bandwidth of 2 days.**

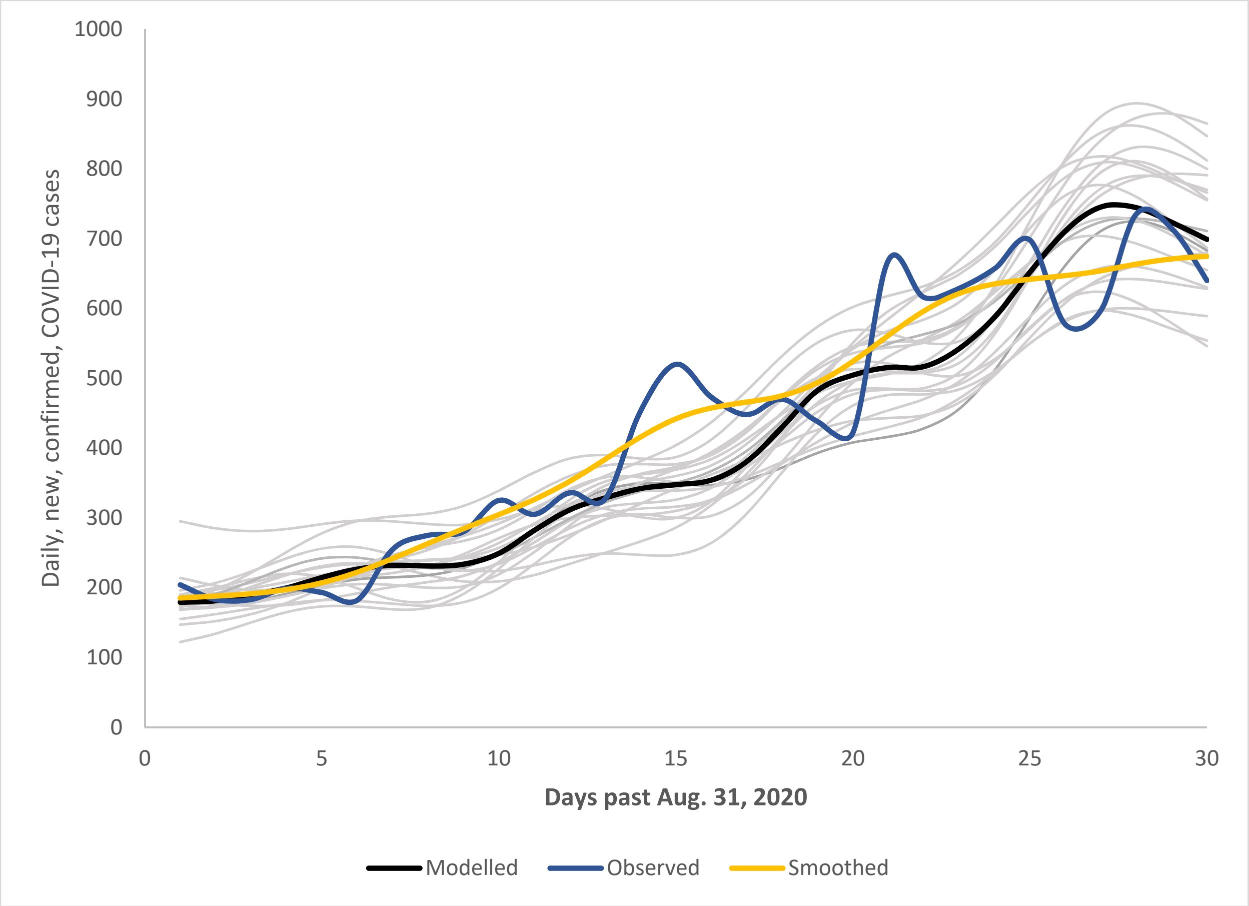


**eFigure 10. Model calibration to daily new, total, confirmed COVID-19 cases in Ontario – second wave.** Observed cases in the CCMPLUS database (blue), after Gaussian kernel smoothing with a bandwidth of 2 days (gold), compared to the average among 20 model replicates (black) and to the individual calibrated replicates (grey) along between September 1 and 30, 2020. Note, both observed and modelled counts subjected to Gaussian kernel smoothing with a bandwidth of 2 days.

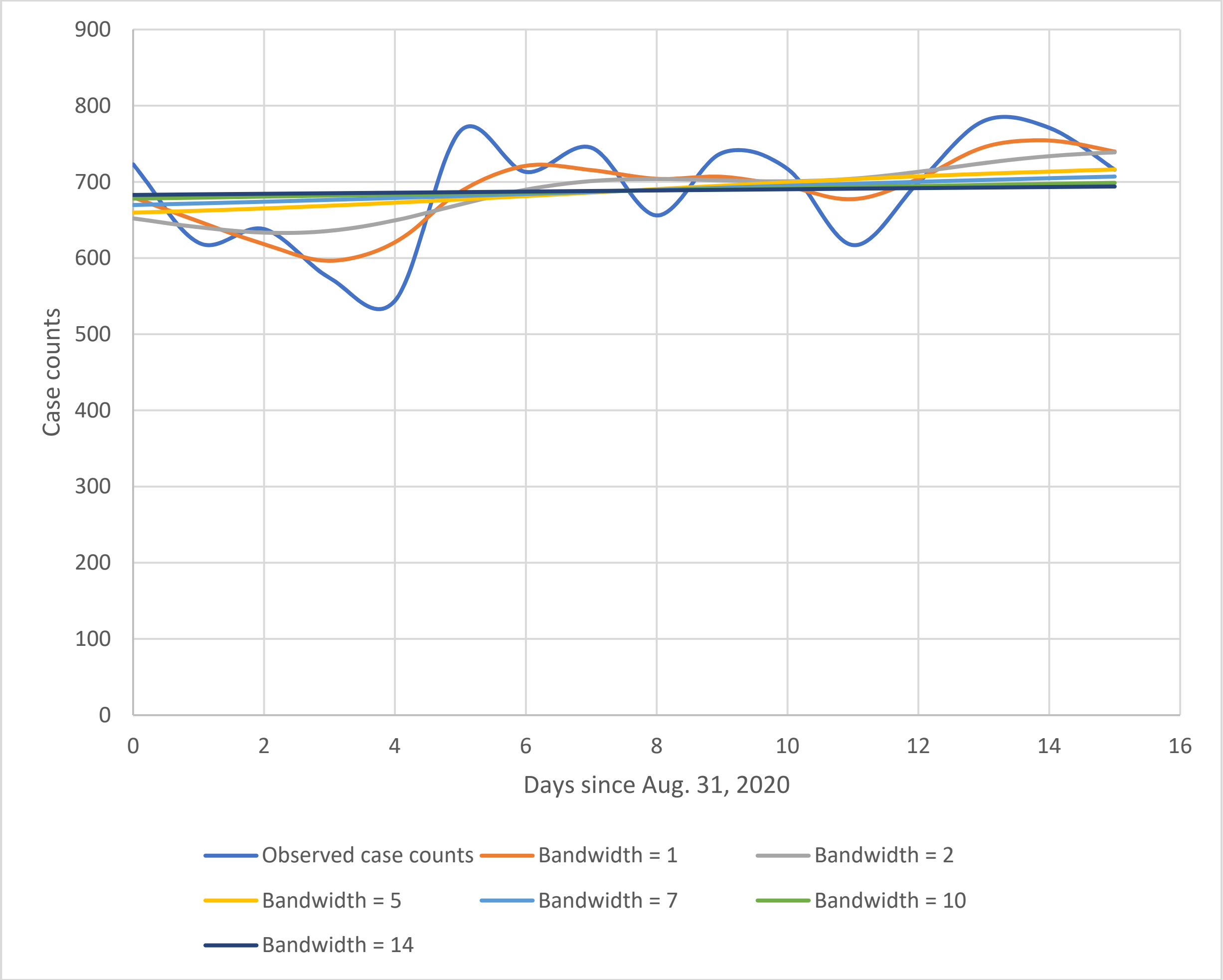


**eFigure 11. Observed versus smoothed daily, new, symptomatic, COVID-19 case counts – during the first 15 days of October, 2020 – for various Gaussian kernel smoothing bandwidths.**

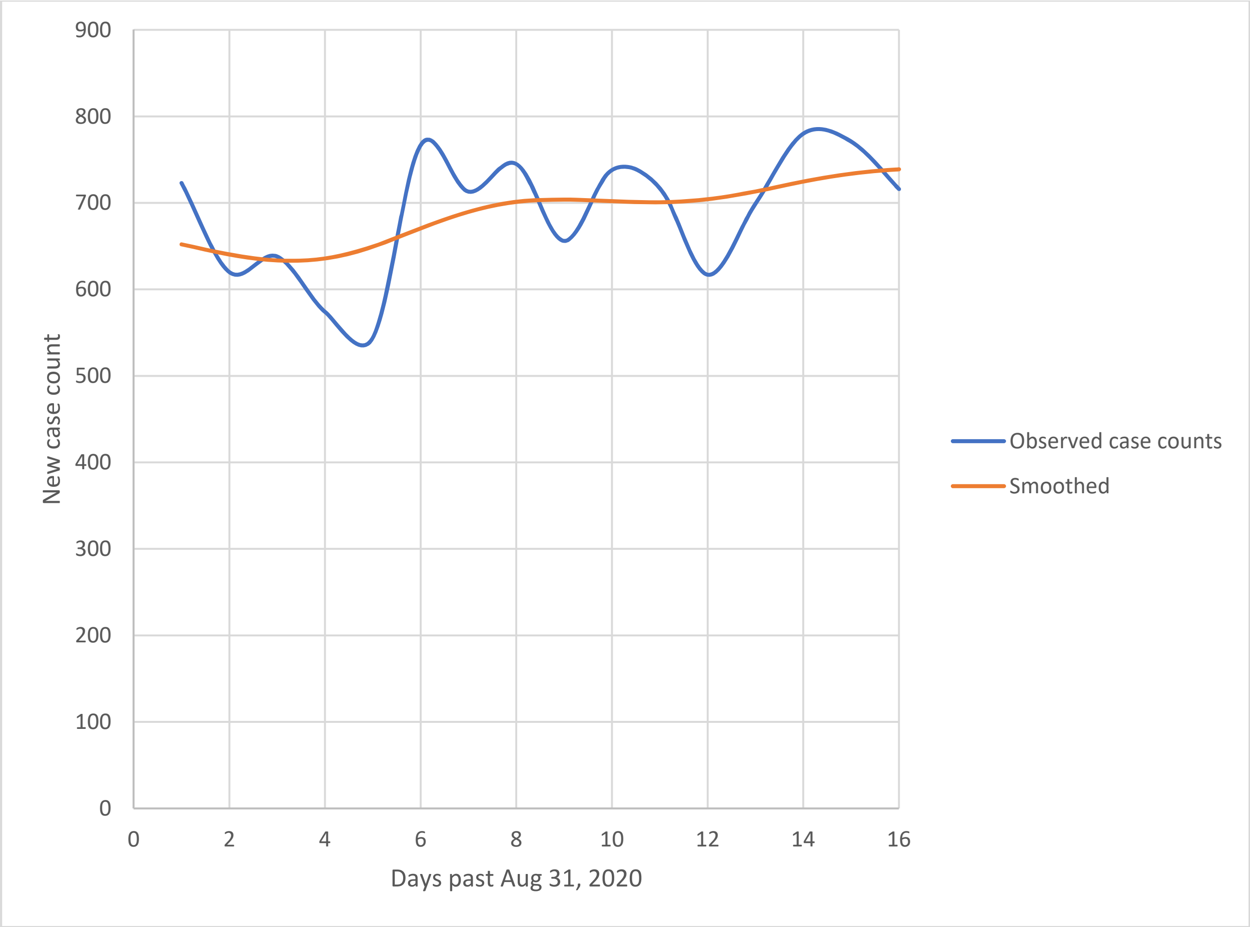


**eFigure 12. Observed versus smoothed daily, new, symptomatic, COVID-19 case counts – during the first 15 days of October, 2020 – for the selected Gaussian kernel smoothing bandwidth of 2 days.**

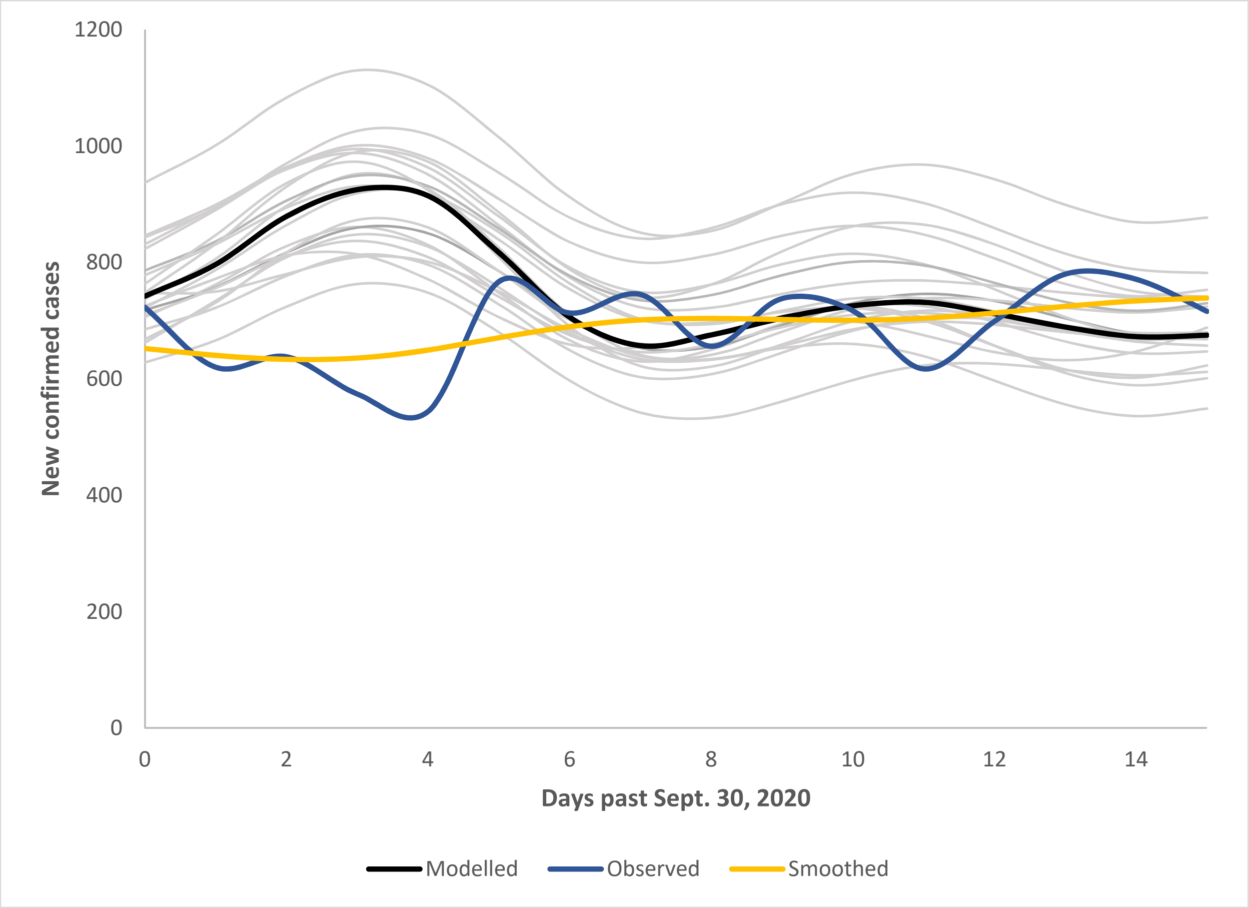


**eFigure 13. Model calibration to daily new, total, confirmed COVID-19 cases in Ontario – first 15 days of October, 2020.** Observed cases in the CCMPLUS database (blue), after Gaussian kernel smoothing with a bandwidth of 2 days (gold), compared to the average among 20 model replicates (black) and to the individual calibrated replicates (grey) along between September 1 and 30, 2020. Note, both observed and modelled counts subjected to Gaussian kernel smoothing with a bandwidth of 2 days.

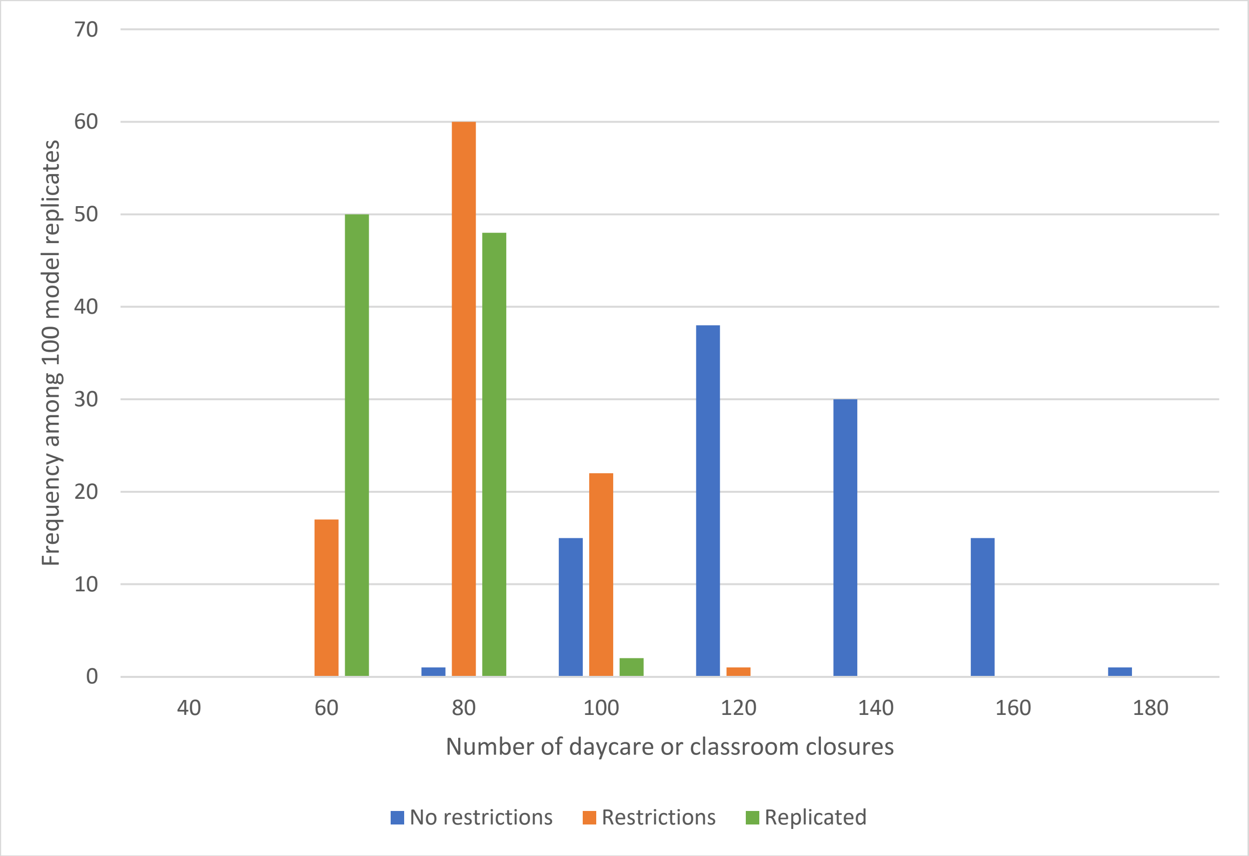


**eFigure 14. Distribution of numbers of daycare, primary, elementary and high school classroom closures for scenarios in which schools had re-opened**: distributions of numbers of classroom closures on model scale among 100 simulation model replicates. Scenario 1b - for the case that no restrictions were implemented (blue); scenario 2b – for the case that restrictions were implemented (orange); scenario 3b - the slowing of the growth of new case numbers observed from October 1 – 15, 2020, had persisted until October 31, 2020 (green).

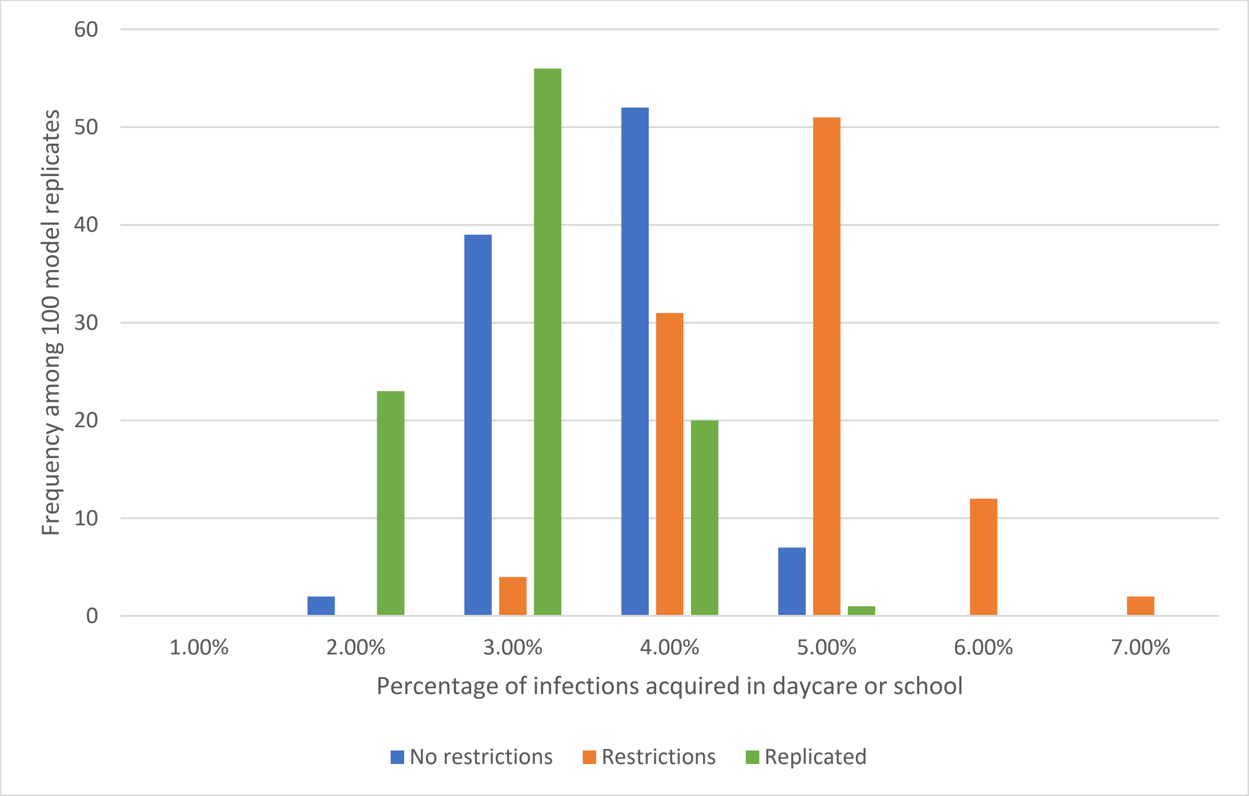


**eFigure 15. Distribution of the percentage of COVID-19 infections acquired in schools for students and teachers in scenarios in which schools had re-opened:** distributions of the percentage of COVID-19 infections acquired in school among the total number of infections among students and teachers on model scale among 100 simulation model replicates. Scenario 1b - for the case that no restrictions were implemented (blue); scenario 2b – for the case that restrictions were implemented (orange); scenario 3b - the slowing of the growth of new case numbers observed from October 1 – 15, 2020, had persisted until October 31, 2020 (green).

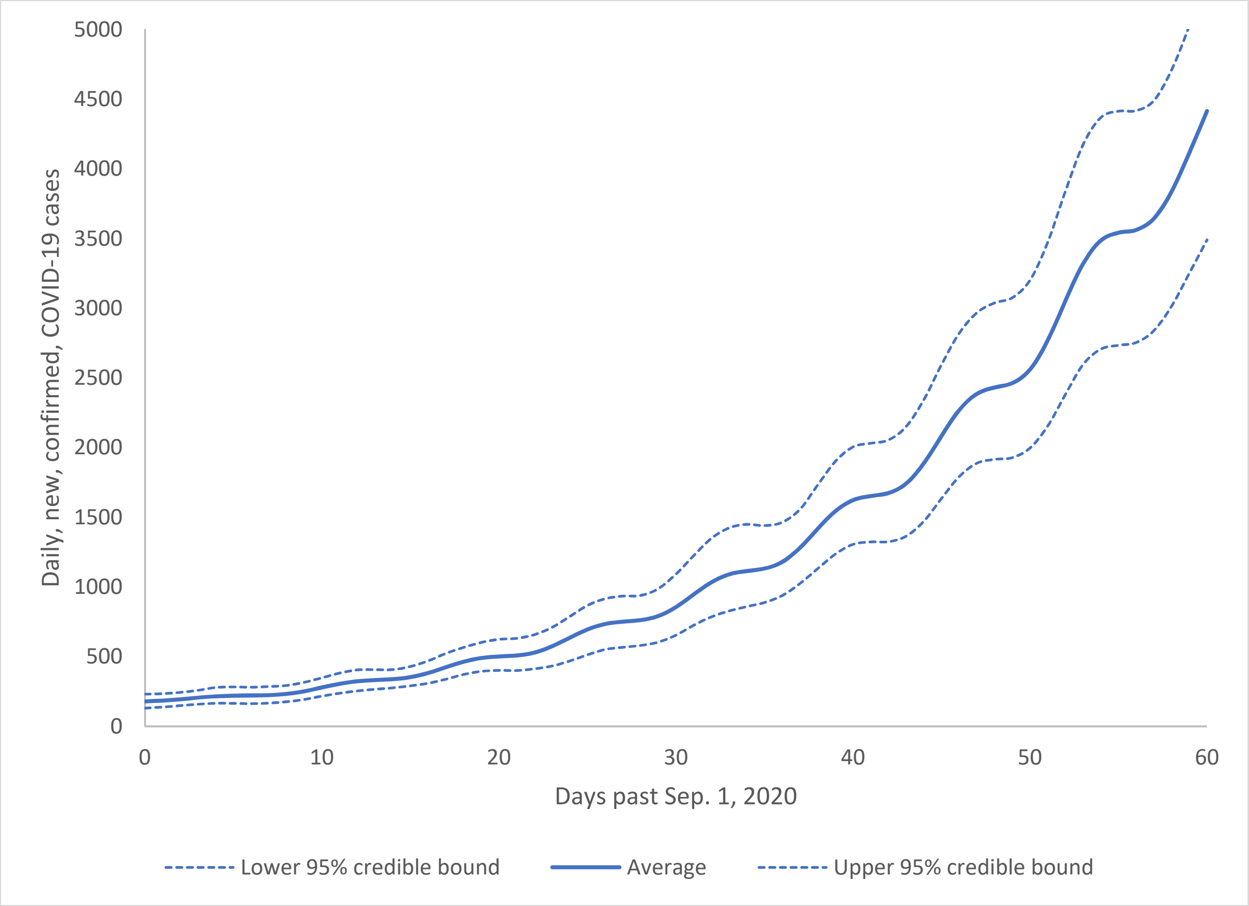


**eFigure 16. Model simulation results for scenario 1a:** no nonpharmaceutical interventions (NPIs) restricting contacts and/or reducing the probability of transmission of COVID-19 between contacts had been implemented (that is, the trends in the rise of new cases in September *had been allowed* to persist through October) and schools *remained closed* on September 15, 2020. Average of 100 model replicates (solid blue) along with the associated 2.5$^{th}$ and 97.5$^{th}$ percentiles (dotted blue) between September 1 and October 31, 2020. Note, modelled counts subjected to Gaussian kernel smoothing with a bandwidth of 2 days.

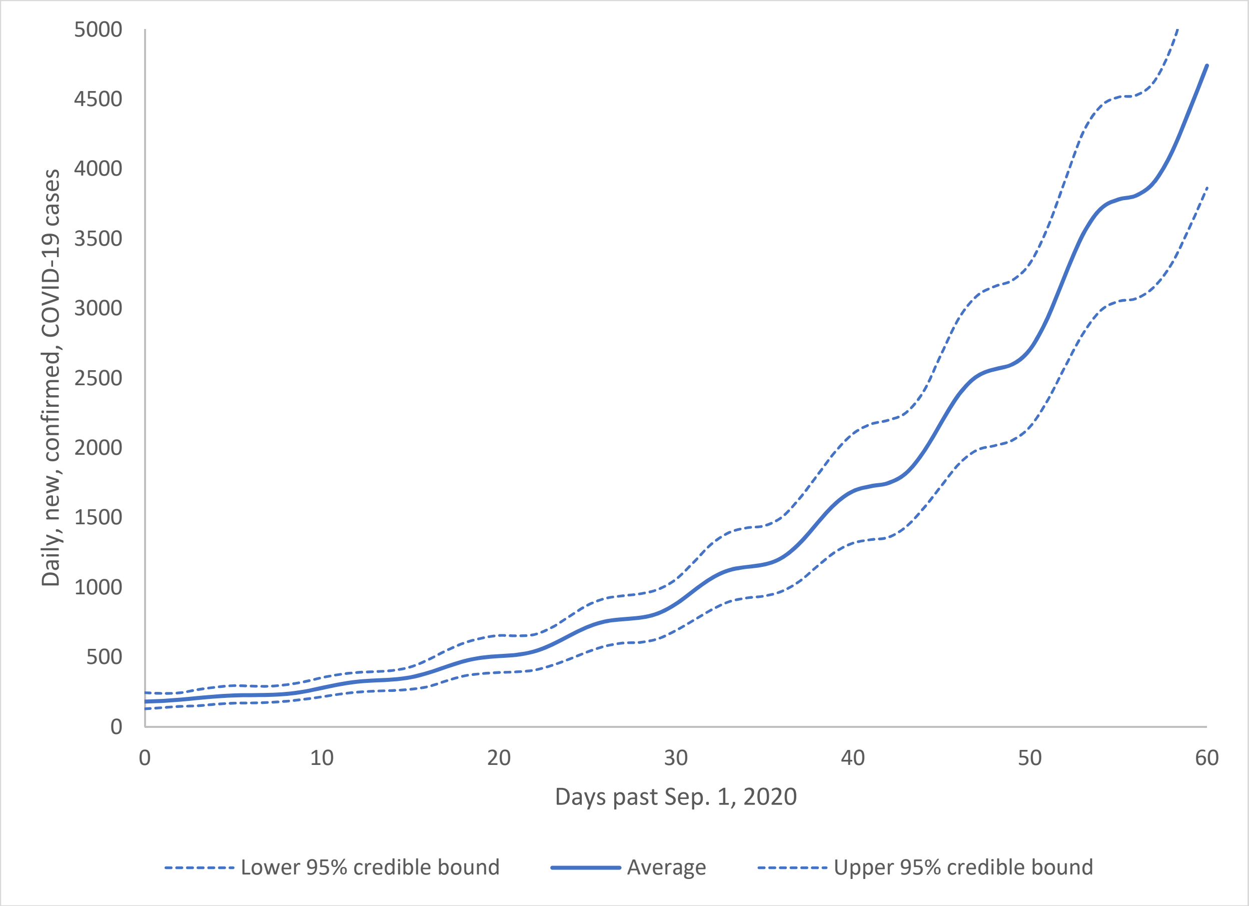


**eFigure 17. Model simulation results for scenario 1b:** no nonpharmaceutical interventions (NPIs) restricting contacts and/or reducing the probability of transmission of COVID-19 between contacts had been implemented (that is, if the trends in the rise of new cases in September *had been allowed* to persist through October) and schools *had re-opened* on September 15, 2020. Average of 100 model replicates (solid blue) along with the associated 2.5$^{th}$ and 97.5$^{th}$ percentiles (dotted blue) between September 1 and October 31, 2020. Note, modelled counts subjected to Gaussian kernel smoothing with a bandwidth of 2 days.

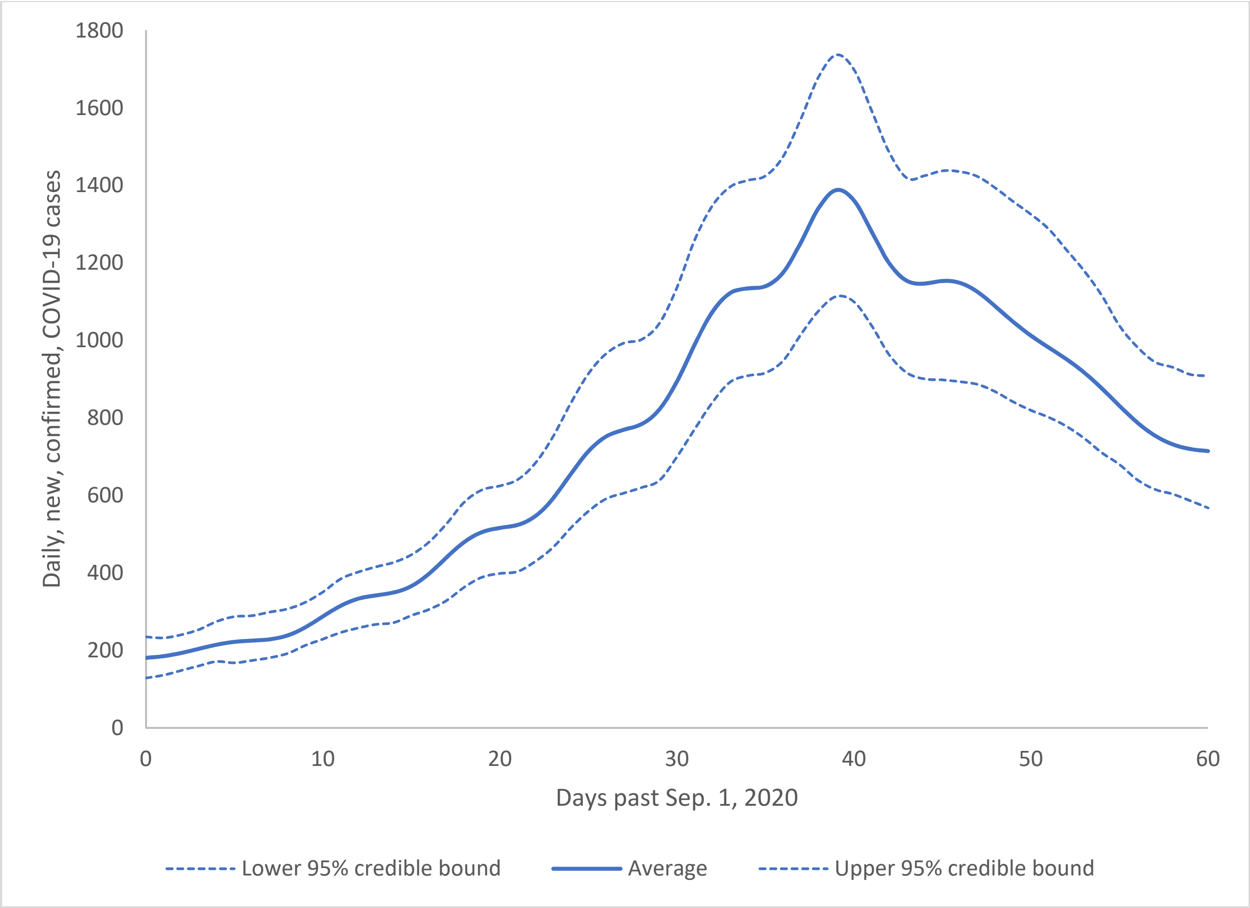


**eFigure 18. Model simulation results for scenario 2a:** nonpharmaceutical interventions (NPIs) restricting contacts and/or reducing the probability of transmission of COVID-19 between contacts had been implemented on October 1, 2020 (that is, if the trends in the rise of new cases in September *had not been allowed* to persist through October) and schools *remained closed* on September 15, 2020. Average of 100 model replicates (solid blue) along with the associated 2.5th and 97.5th percentiles (dotted blue) between September 1 and October 31, 2020. Note, counts subjected to Gaussian kernel smoothing with a bandwidth of 2 days.

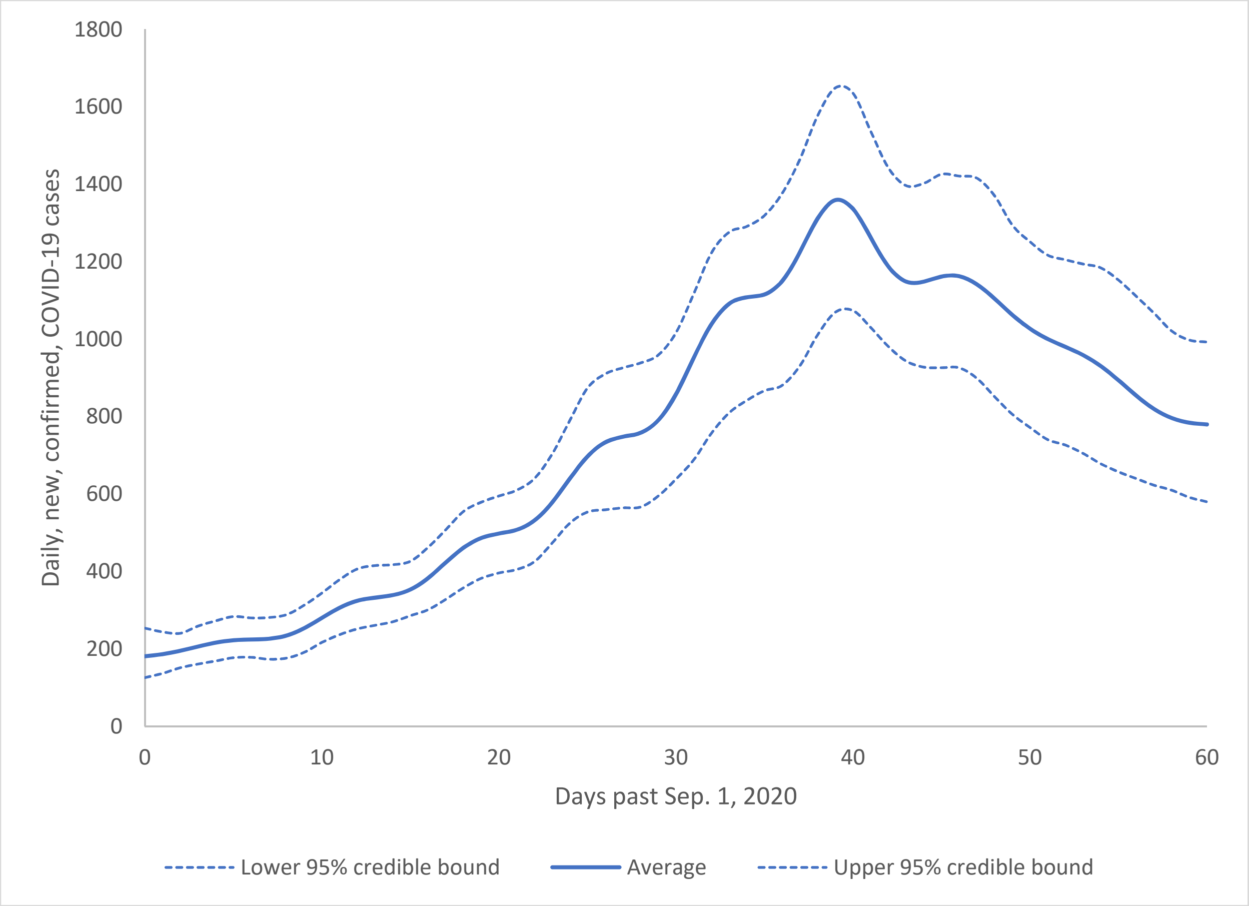


**eFigure 19. Model simulation results for scenario 2b:** nonpharmaceutical interventions (NPIs) restricting contacts and/or reducing the probability of transmission of COVID-19 between contacts were implemented on October 1, 2020 (that is, if the trends in the rise of new cases in September *had not been allowed* to persist through October) and schools *had re-opened* on September 15, 2020. Average of 100 model replicates (solid blue) along with the associated 2.5$^{th}$ and 97.5$^{th}$ percentiles (dotted blue) between September 1 and October 31, 2020. Note, counts subjected to Gaussian kernel smoothing with a bandwidth of 2 days.

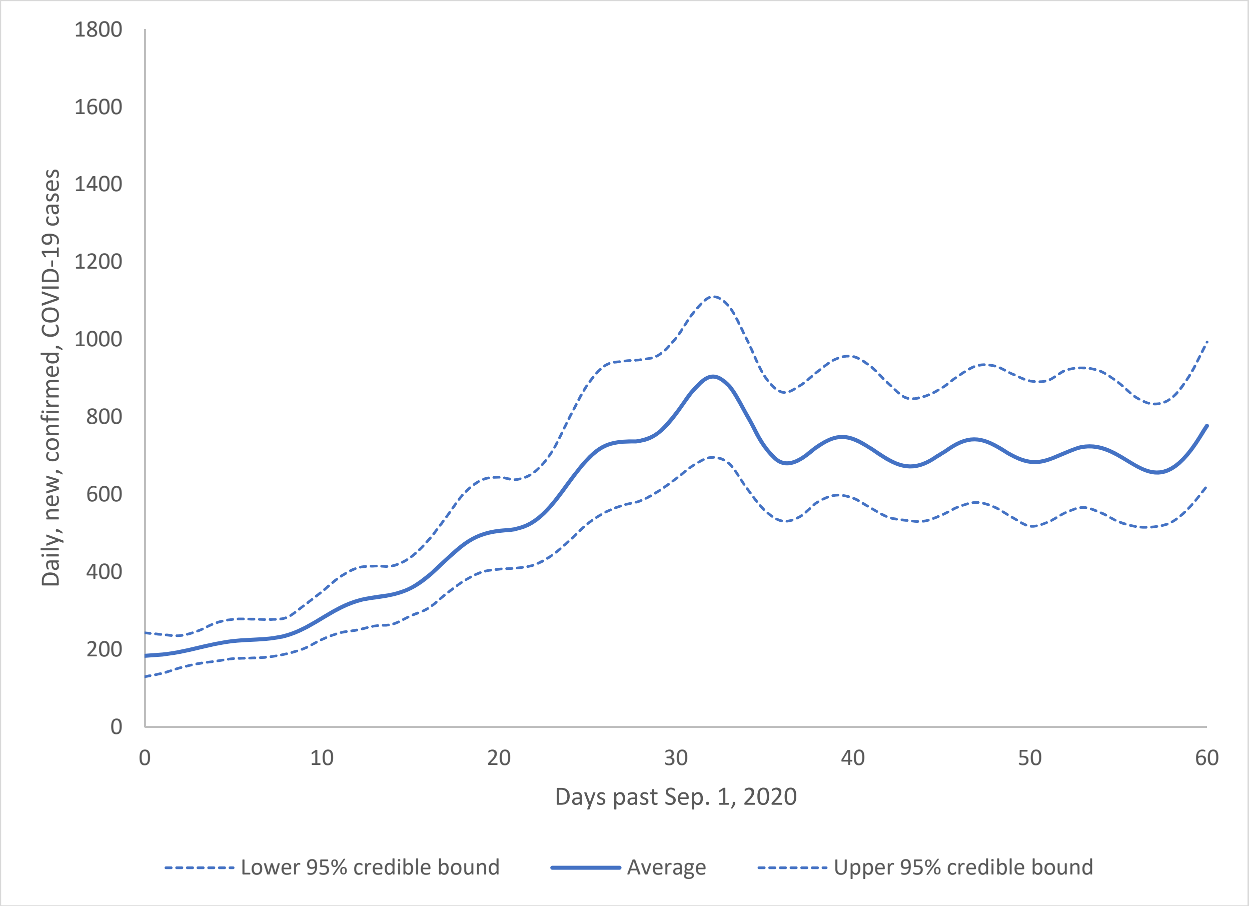


**eFigure 20. Model simulation results for scenario 3a:** the reduction of the growth in new, daily, confirmed, COVID-19 cases had reduced to that observed between October 1 – 15, 2020, the lower growth rate had persisted until Oct 31, 2020, and schools *remained closed* on September 15, 2020. Average of 100 model replicates (solid blue) along with the associated 2.5$^{th}$ and 97.5$^{th}$ percentiles (dotted blue) between September 1 and October 31, 2020. Note, counts subjected to Gaussian kernel smoothing with a bandwidth of 2 days.

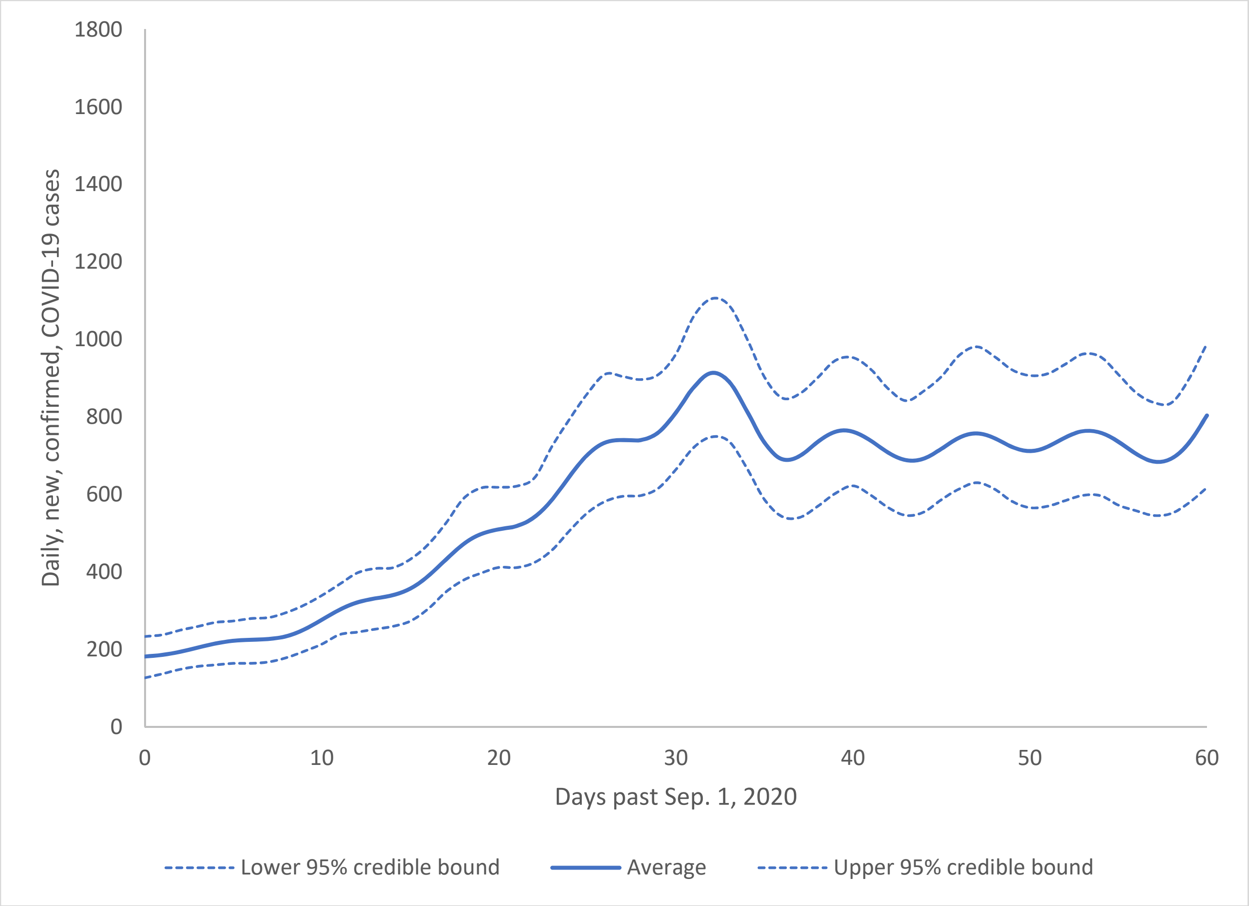


**eFigure 21. Model simulation results for scenario 3b:** the reduction of the growth in new, daily, confirmed, COVID-19 cases had reduced to that observed between October 1 – 15, 2020, that the lower growth rate had persisted until Oct 31, 2020, and schools *had re-opened* on September 15, 2020. Average of 100 model replicates (solid blue) along with the associated 2.5$^{th}$ and 97.5$^{th}$ percentiles (dotted blue) between September 1 and October 31, 2020. Note, counts subjected to Gaussian kernel smoothing with a bandwidth of 2 days.

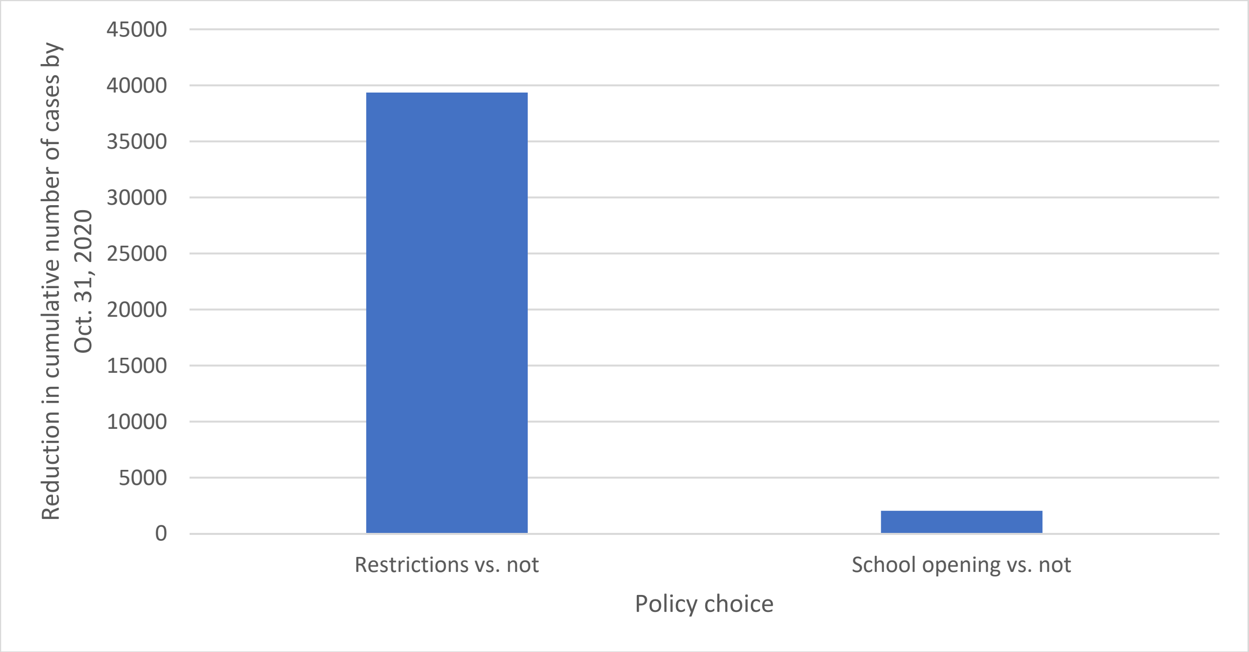


**eFigure 22. Reduction of cumulative COVID-19 cases between September 1 and October 31, 2020, attributable to the two policy choices**: to implement nonpharmaceutical interventions (NPIs) or not (the average of the cumulative cases for scenarios 1a and 1b minus the average for scenarios 2a and 2b) vs. to open schools or not (the average for scenarios 1b and 2b minus the average for scenarios 1a and 2a).

**eAppendix References**


1. Statistics Canada. Social Policy Simulation Database and Model (SPSD/M) Version 28.0.1. 2020. https://search2.odesi.ca/#/details?uri=%2Fodesi%2Fspsd-89F0002-E.xml.
2. Statistics Canada. *Rural and Small Town Canada Analysis Bulletin (Cat.21-006-XIE).*; 2001.
3. Ontario Ministry of Health. *Contact and Case Management System (CCMPLUS).*; 2020.
4. Tuite AR, Fisman DN, Greer AL. Mathematical modelling of COVID-19 transmission and mitigation strategies in the population of Ontario, Canada. *Cmaj*. 2020;192(19):E497-E505. doi:10.1503/cmaj.200476
5. Ontario Public Health. *COVID-19 Seroprevalence in Ontario: March 27, 2020 to June 30, 2020.*; 2020.
6. Brisson, Marc; Gingras, Guillaume; Drolet, Melanie; Laprise J-F. Modélisation de l'évolution de la COVID-19 au Québec. http://www.marc-brisson.net/covid19-response/Epidemiologie-et-modelisation-evolution-COVID-19-au-Quebec_16-octobre-2020.pdf.
7. Statistics Canada. *Percentage of Workforce Teleworking or Working Remotely, and Percentage of Workforce Expected to Continue Teleworking or Working Remotely after the Pandemic, by Business Characteristics*.
8. Powell M. *The BOBYQA Algorithm for Bound Constrained Optimization without Derivatives*. Cambridge; 2010. http://www.optimization-online.org/DB_HTML/2010/05/2616.html.
9. Wang, S; Kustra, R; Watts, A; Khan K; Mishra S. *GTA Proxy Measures of Physical Distancing.*; 2020.
